# Immunogenicity and vaccine effectiveness of COVID-19 vaccines in people on immunosuppressive therapies

**DOI:** 10.64898/2026.09.16.26363194

**Authors:** Eva Stadler, Shanchita R Khan, Karen M Elias, Ece Egilmezer, Chansavath Phetsouphanh, Priyanka Hastak, Rehana V Hewavisenti, Ruchika V Joshi, Tari Turner, Deborah Cromer, Sarah C Sasson, Miles P Davenport, David S Khoury

## Abstract

Neutralizing antibodies are a correlate of protection (CoP) of SARS-CoV-2 vaccine efficacy. However, data informing CoPs often explicitly exclude people on immunosuppressive therapies who are at an increased risk of symptomatic and severe COVID-19, delayed viral clearance, and death. Investigating the relationship between antibodies and protection for people on immunosuppressive therapies could provide insights into mechanisms of protection.

We performed a systematic search to identify studies reporting SARS-CoV-2 binding antibody levels after mRNA vaccination in people with hematological malignancies, neurological disorders, rheumatic diseases, and mixed immune-mediated inflammatory disorders (IMIDs) on immunosuppressive therapies including B-cell depletion, JAK inhibitors, and S1P inhibitors, as well as healthy controls. Binding antibody levels varied between cohorts receiving different immunosuppressive treatments. Compared to healthy controls the lowest antibody levels were observed in subjects treated with B-cell depletion (54.5-fold reduction, 95% CI: 34.5-86.2) and S1P inhibitors (24.8-fold reduction, 95% CI: 13.2-46.7).

To assess the association between antibody level and protection, we used data from a previous study of COVID-19 vaccine effectiveness in people with the same underlying conditions. We linked vaccine effectiveness estimates with the predicted antibody level in people on immunosuppressive therapies (using antibody data including this meta-analysis of binding antibody levels). Predicted neutralizing antibody levels were correlated with vaccine effectiveness both for infection and hospitalization (p<0.0001 and p<0.0001, respectively). However, we found that for any given antibody level, people who are immunosuppressed had lower protection against infection and hospitalization than a healthy population.

## Introduction

Both neutralizing and binding antibodies are correlated with protection from SARS-CoV-2 in the general (immunocompetent) population^1–4^. Studies of vaccine responses in subjects undergoing immunosuppressive treatments have shown reduced antibody levels and reduced vaccine effectiveness^5, 6^. By contrast, T cell responses to vaccination are often relatively preserved in people on some immunosuppressive therapies^7, 8^, and studies have suggested that these responses may maintain protection from severe COVID-19^9^.

Neutralizing antibody correlates of protection (CoPs) for COVID-19 are used for public health decision-making, including the approval of new vaccines and therapeutics^10–13^. People on immunosuppressive therapies are often excluded from vaccine trials^14^, and it is unclear if antibodies may also serve as a correlate of protection from COVID-19 in these populations. Despite this uncertainty, studies of antibody levels after vaccination in immunosuppressed populations are commonly used to guide therapy for this at-risk population^7, 15^. Therefore, it is critical to evaluate whether antibodies may also be associated with vaccine effectiveness in people on immunosuppressive therapies. Studies of vaccine immunogenicity and effectiveness in people receiving immunosuppressive therapies provide an opportunity to assess the relevance of established CoPs for COVID-19 in this population.

Here, we performed a systematic search to identify studies of binding antibody levels after mRNA vaccination in people on immunosuppressive therapy, including patients with underlying hematological malignancies and autoimmune and inflammatory disorders. We linked these data with data on vaccine effectiveness from our previous systematic review in people on immunosuppressive therapies^16^ to test whether there is evidence of an association between antibody levels and vaccine effectiveness in people on immunosuppressive therapies.

## Results

### Systematic search for studies reporting binding Ab levels after vaccination

To assess how different immunosuppressive treatments impact antibody (Ab) levels after vaccination, we conducted a systematic search of studies of antibody binding in people with secondary immunosuppression. We chose to focus on binding antibody levels following mRNA vaccinations in people who are immunosuppressed because preliminary scoping searches indicated substantially more data was available on binding levels than on neutralizing antibody levels in these cohorts, as well as more data for mRNA vaccines than other COVID-19 vaccine platforms. To investigate the relative impact of different underlying conditions and immunosuppressive therapies on Ab levels, we chose high-prevalence conditions that are often managed with a common set of immune-modulating interventions (Materials and Methods). The full details of search, screening, extraction, inclusion and exclusion criteria are reported in the Supplement (pp. 3-5, Table S1).

Our search returned 2,656 articles after removing duplicates. Following screening, we identified 19 studies meeting our inclusion criteria (Fig. S1, Table S1)^17–35^. Studies reported binding Ab levels in people with hematologic malignancies (Haem, n=5 of 19 studies, 26%)^20, 23, 24, 31, 33^, mixed immune-mediated inflammatory disorders (IMIDs, n=3, 16%)^21, 29, 34^, neurological disorders (Neur, n=7, 37%)^18, 19, 25, 27, 28, 30, 32^, and rheumatic diseases (Rheu, n=4, 21%)^17, 22, 26, 35^ (Table S1-2). We grouped immunosuppressive treatments into seven categories: 3 (16%) studies reported on people receiving anti-IL6R therapy^17, 22, 35^, 5 (26%) studies for anti-TNFα therapy^17, 22, 26, 34, 35^, 11 (58%) studies for B-cell depletion treatments^17–19, 21, 22, 27, 28, 30–33^, 4 (21%) studies for treatment with csDMARDs (conventional synthetic Disease Modifying Anti-Rheumatic Drugs) ^22, 26, 29, 35^, 4 (21%) studies of integrin inhibitors^25, 27, 28, 34^, 8 (42%) studies of JAKi (JAK inhibitors)^17, 20, 22–24, 26, 34, 35^, and 7 (37%) studies of S1P inhibitors (sphingosine-1-phosphate inhibitors)^18, 19, 25, 27, 28, 30, 32^. Additionally, four studies included people with the same conditions who were not receiving immunosuppressive therapies^28, 30, 31, 34^. The vaccines used were Pfizer (n=9 of 19 studies, 47%)^17, 18, 21, 22, 25, 27, 28, 31, 33^, Moderna (n=3, 16%)^23, 30, 33^, or a mix of these vaccines (n=10, 53%)^19, 20, 24–26, 29, 30, 32, 34, 35^. All studies reported Ab binding levels after two and three doses of an mRNA vaccine. Antibody levels after vaccination were reported as geometric mean antibody binding concentrations or endpoint dilution titers and are referred to as “antibody level” for simplicity.

### Different immunosuppressive treatments are associated with different effects on vaccine immunogenicity

Comparing across all included conditions and treatments, we found that the people who are immunosuppressed had, on average, 6.4-fold lower antibody (Ab) levels than healthy controls (95% confidence interval, CI: 4.0-10.2, Table S3). For each underlying condition, at least three different immunosuppressive therapies were used in the included studies and each immunosuppressive therapy was used across at least two different underlying condition categories – except S1P inhibitors used only to treat neurological disorders and anti-IL6R used only to treat rheumatic diseases (Fig. S2). Therefore, we next considered whether antibody levels after vaccination were influenced by the underlying condition (for which immunosuppressive treatment was administered) or by the immunosuppressive treatment itself. To aid this analysis, we also included data from four studies that enrolled groups of people with underlying conditions that typically require immunosuppressive therapies but who were not on immunosuppressive treatment during the study^28, 30, 31, 34^. We first noted that both underlying condition and immunosuppressive treatment were significantly associated with antibody binding levels after vaccination (multivariate model included the number of vaccinations and time since last vaccination and either condition or treatment; condition: p<0.0001, treatment: p<0.0001, Table S4, **Fig. 1**, **Fig. 2**). We found that if immunosuppressive treatment type was included as a predictor of log_10_-Ab levels, then adding the underlying condition as a predictor did not improve the model’s predictive value (as determined by likelihood ratio test, p=0.26, Table S4). However, for a model using condition as a predictor, adding treatment type as an additional predictor significantly improved the model fit (likelihood ratio test, p<0.0001, Table S4, Table S5). Overall, model selection revealed that the best model (by Akaike Information Criterion with small sample size correction, AICc) included the number of vaccine doses received, time since last vaccination and immunosuppressive treatment type but it did not include underlying condition (Table S6, **Fig. 1**A-B, Fig. S3). Examining this further, we found that B-cell depletion and S1P inhibitor treatments led to significantly lower Ab levels than all other treatments, with 54.5-fold (95% CI: 34.5-86.2) and 24.8-fold (95% CI: 13.2-46.7) lower Ab levels, respectively, compared to no immunosuppressive treatment (**Fig. 1**C, Table S7). Of the other treatment types, JAKi were associated with 3.8-fold lower Ab levels, anti-IL6R with 3.3-fold lower Ab levels, anti-TNFα with 2.6-fold lower Ab levels, csDMARDs with 1.6-fold lower Ab levels, and integrin inhibitors with 1.2-fold lower Ab levels compared to no immunosuppressive treatment (all differences were significant except for csDMARDs and integrin inhibitors, contrast analysis; **Fig. 1**C, Table S7). Together, our findings show that Ab levels following vaccination are significantly lower for some immunosuppressive therapies and also vary across underlying conditions, with stronger evidence for differences driven by immunosuppressive treatment than by underlying condition (**Fig. 1**, **Fig. 2**, Table S4-8).

**Fig. 1.**
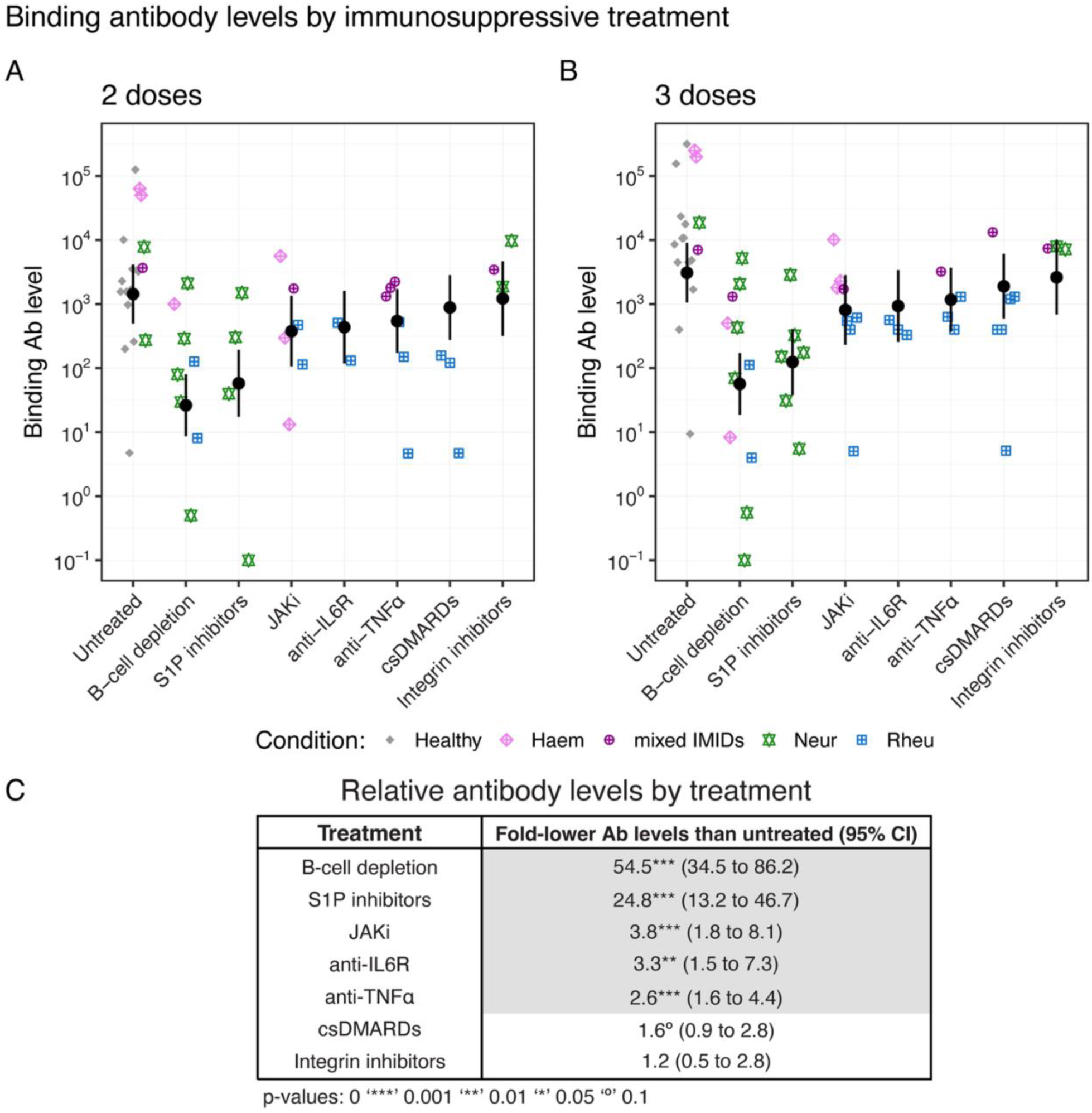
Binding antibody levels in people on immunosuppressive therapies by immunosuppressive treatments for two and three doses of an mRNA vaccination within two months after vaccination. (A, B) Binding Ab levels are grouped by treatment with underlying conditions indicated by different colors and shapes, and black points show the Ab level estimates using a mixed-effects model (with 95% confidence bounds shown as vertical black lines, the statistical model includes treatment, number of vaccinations (dose), and time since last vaccination as predictors; Table S4, model 3). (C) Difference in binding Ab level for immunosuppressive treatments compared to untreated individuals with p-values for comparison of each treatment with untreated individuals using pairwise contrast analysis (significant difference, p < 0.05, is indicated by grey shading, Table S7). Comparisons show significantly lower Ab levels for patients receiving B-cell depletion or S1P inhibitor treatments compared to other immunosuppressive regimens (csDMARDs, anti-IL6R, integrin inhibitors, JAKi, and anti-TNFα) with 54.5- and 24.8-fold lower levels than untreated patients, ectively. A sensitivity analysis excluding censored Ab levels also showed significantly lower Ab levels for B-cell depletion and S1P inhibitor treatments (Table S5-6, Table S8).

**Fig. 2.**
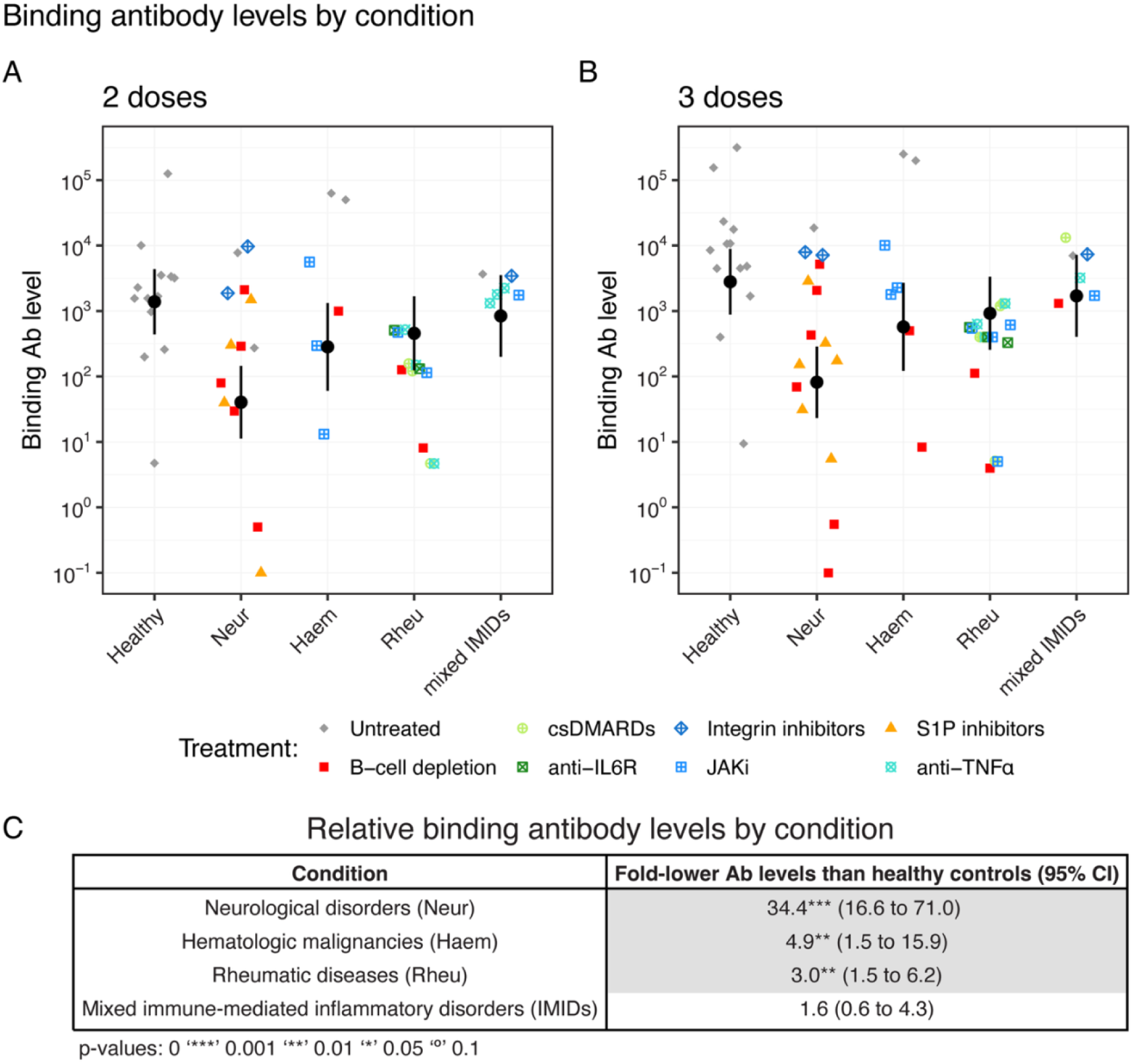
Binding antibody levels in people on immunosuppressive therapies by underlying conditions for two and three doses of an mRNA vaccination within two months after vaccination. (A, B) Binding Ab levels are grouped by condition with immunosuppressive treatments indicated by different colors and shapes, and black points show the Ab level estimates using a mixed-effects model (with 95% confidence bounds shown as vertical black lines, the statistical model includes condition, doses, and time since last vaccination as predictors; Table S4, model 4). (C) Difference in binding Ab level for underlying conditions compared to healthy controls with p-values for comparison of each condition with healthy controls using pairwise contrast analysis (significant difference, p < 0.05, is indicated by grey shading, Table S7). Comparisons show significantly lower Ab levels for patients with neurological disorders (Neur), hematologic malignancies (Haem), and rheumatic diseases (Rheu) compared to healthy controls but no significant Ab level reduction for people with mixed immune-mediated inflammatory disorders (IMIDs). A sensitivity analysis excluding censored Ab levels also showed significantly lower Ab levels for people with neurological disorders, hematologic malignancies, and rheumatic diseases (Table S5, Table S8).

### Ab level and vaccine effectiveness for people on immunosuppressive therapies

Both neutralizing and binding antibody levels have been shown to be predictors of protection against COVID-19 in a general population and neutralizing antibody titers continue to be predictive of protection against SARS-CoV-2 variants^1, 4, 36, 37^. We sought to determine whether antibody levels were also associated with vaccine effectiveness (VE) in people on immunosuppressive therapies. We combined the immunogenicity data identified in this work with effectiveness data previously reported in a systematic review and meta-analysis of vaccine effectiveness in people on immunosuppressive therapies with the same conditions (Table S9)^16^. One challenge is that the VE estimates varied by vaccine used, number of vaccine doses, time since vaccination, and circulating SARS-CoV-2 variants. To match the data from immunogenicity and protection studies, we adapted an approach previously used in healthy subjects^36^. This involved estimating antibody responses to specific variants after a given number of vaccine doses using data from the Stanford University Coronavirus Antiviral & Resistance Database^38^ (Standford Data Base, SDB) and adjusting for the additional effects of immunosuppressive treatment or underlying condition (Materials and methods, Supplement pp. 8-9). Specifically, we matched each reported VE estimate to a predicted neutralizing Ab level based on the neutralizing Ab titer from Khoury et al. (2021)^1^ for the same vaccine with adjustments for doses and circulating variant using the SDB and for the fold-drop in antibody binding for the underlying condition or immunosuppressive treatment as estimated in this study (Fig. S4).

We found that the predicted log_10_-antibody levels were significantly associated with VE for each outcome (p<0.0001, p<0.0001, and p=0.0075 for the clinical outcomes of infection, hospitalization, and death, respectively, Table S10, **Fig. 3**, Fig. S5). These results suggest that for people on immunosuppressive therapies included in our analysis, predicted antibody levels (adjusted by loss of neutralization to different variants) are correlated with protection against SARS-CoV-2 infection, hospitalization, and death.

**Fig. 3.**
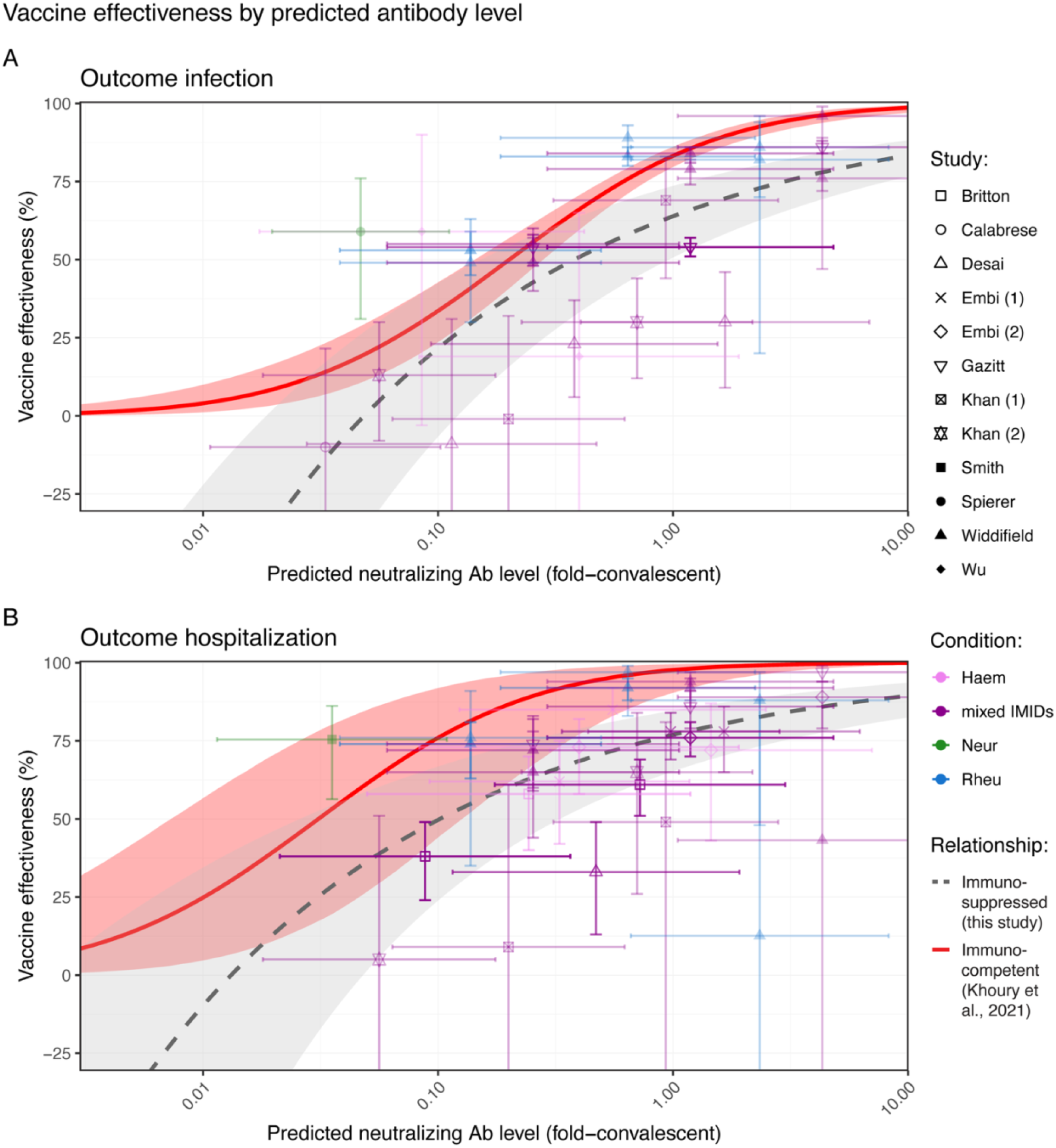
Comparison of the relationship between vaccine effectiveness (VE) and predicted neutralizing antibody (Ab) levels in people on immunosuppressive therapies (grey dashed line with shaded 95% confidence region) and immunocompetent (Khoury et al. (2021)^1^, red line, with red-shaded 95% confidence region) for outcomes infection (A) and hospitalization (B). Predicted Ab levels were linked based on the underlying condition or immunosuppressive treatment (Supplement pp. 8-9, Fig. S4). The fit to the data includes log10-Ab levels as the only predictor and a random intercept by study (Table S10). The opacity of the data is based on the inverse variance of the vaccine effectiveness estimate. For outcome death see Fig. S5. A sensitivity analysis excluding censored Ab levels showed consistent results (Fig. S8, Table S10-11, Table S13).

### Comparing correlates of protection for people on immunosuppressive therapies and immunocompetent individuals

Finally, we considered whether the relationship between Ab levels and protection observed here is similar or different to the relationship we identified previously from a meta-analysis of vaccine efficacy in the general population^1^. For SARS-CoV-2 infection, the association between predicted neutralizing antibody level and vaccine protection was similar, however the estimated protection was lower (for equivalent Ab levels) than the previously reported association in immunocompetent individuals (**Fig. 3**A). We also observed high heterogeneity between studies (I^2^ 94.1% and 86.9% for outcomes infection and hospitalization, respectively, Table S10). Similarly, the relationship between predicted neutralizing Ab levels and vaccine effectiveness against hospitalization in people on immunosuppressive therapies showed a lower protection than that for the general population at the same Ab level, with most (29/31, 93.5%) vaccine effectiveness estimates in people on immunosuppressive therapies being below the expected vaccine effectiveness predicted by a previously derived association for the general population (**Fig. 3**B). Together this suggests that people on immunosuppressive therapies have a lower vaccine-induced protection compared to immunocompetent individuals, even at the same predicted antibody level.

## Discussion

We identified and analyzed binding Ab data following mRNA vaccination in people with hematological malignancies, neurological disorders, rheumatic diseases, and mixed immune-mediated inflammatory disorders. Antibody levels differed significantly between conditions, with the lowest Ab levels in people with neurological disorders, likely related to the high use of B-cell depletion and S1P inhibitors (**Fig. 2**). We also considered eight different treatment types and found that people on B-cell depletion or S1P inhibitor treatments had the lowest Ab levels (**Fig. 1**). JAKi, anti-IL6R, and anti-TNFα treatments were also associated with significantly lower Ab levels whereas we found only a small reduction in Ab levels (not statistically significant) in individuals receiving csDMARDs or integrin inhibitor treatments (**Fig. 1**). Statistical testing suggested that immunosuppressive therapy was a bigger determinant of binding Ab levels after vaccination than the underlying condition (Table S4-5).

The significantly lower Ab levels in people receiving B-cell depletion and S1P inhibitor treatments compared to other immunosuppressive treatments (csDMARDs, anti-IL6R, integrin inhibitors, JAKi, and anti-TNFα) agrees with previous reports^39–41^. It is interesting to note that in our data, immunosuppressive treatment was more important than underlying condition as a predictor of vaccine-induced binding Ab levels. The finding that csDMARDs and integrin inhibitors were not associated with significant decreases in Ab levels compared to healthy controls (1.6-fold and 1.2-fold lower Ab levels than untreated controls, respectively; p=0.084 and p=0.71, respectively) is also of interest. This may inform clinical decision making in diseases such as inflammatory bowel disease and multiple sclerosis, where integrin inhibitors and csDMARDs have proven efficacy and may be prioritized above B-cell depletion, S1P inhibitors, or TNFα inhibitors in certain settings including during pandemic vaccination rollouts. Overall, this highlights the importance of considering immunosuppressive treatments in decision-making and reporting outcomes, such as vaccine effectiveness, in cohorts including people who are immunosuppressed.

We additionally show that in people on immunosuppressive therapies, predicted Ab levels following vaccination are associated with protection against SARS-CoV-2 infection, hospitalization, and death (Table S10). This suggests that it may be possible to use antibody levels in guiding clinical decision making for people who are immunosuppressed. However, it is important to consider the limitations of the data and their impact on conclusions. For example, this study is based on population-level data rather than individual-level data, combined data across four pre-specified conditions, vaccine effectiveness and Ab level were not reported in the same cohorts, and in the VE data there is very limited information on immunosuppressive therapies (Table S9) and how long after vaccination VE was assessed. Since Ab levels were not directly measured in the clinical efficacy studies, they had to be predicted based on the vaccine, number of vaccinations, circulating variant, and underlying condition or immunosuppressive treatment. Further research is needed to guide clinical decision making in at risk populations such as people on immunosuppressive therapies.

We find that protection against SARS-CoV-2 infection and hospitalization is lower in people on immunosuppressive therapies than in the general population at the same Ab level (**Fig. 3**). This suggests that immunosuppressive treatments and conditions can cause a reduction in vaccine protection beyond what can be accounted for by the reduction of antibody responses and that other immune mechanisms or susceptibility pathways may also contribute protection. Interestingly, B cell depletion treatment causes the greatest reduction in antibody levels but has also been reported to be associated with higher T-cell responses in some patients^9, 42^. Of note, although 29 of 31 vaccine effectiveness estimates against hospitalization in people on immunosuppressive therapies are below the protection predicted in the general population at the same Ab level, the only estimate of vaccine effectiveness against hospitalization that lies well above this line is for a study of people with multiple sclerosis on B-cell depleting therapy (Smith et al.^43^, **Fig. 3**B; the other data point’s vaccine effectiveness 95% CI overlaps with the estimated in the general population, Widdifield et al.^44^, **Fig. 3**B). However, conclusions about mechanisms of protection in B-cell depletion-treated people are speculative and cannot be answered with the data available in this study (this is the only data point for outcome hospitalization and B-cell depletion treatment). Moreover, it is unclear whether (or to what extent) other factors such as behavioral differences between the general population and people on immunosuppressive therapies influence the risk of SARS-CoV-2 infection. For example, people who are immunosuppressed may be more likely to wear masks and shields or practice social distancing compared to the general population, thereby reducing exposure, but they may also be more likely to be required to attend high risk medical settings (GP visits or hospital admissions) which may increase exposure.

This study has several limitations. The vaccine effectiveness data often did not include any information on immunosuppressive treatments or time since last vaccination, and in some cases vaccine effectiveness was not disaggregated by the number of vaccinations^16^. We utilized immunogenicity and effectiveness data following vaccination with vaccines containing the ancestral SARS-CoV-2 immunogen only. This was to ensure alignment with the criteria of the systematic review of vaccine effectiveness in people on immunosuppressive therapies and to reduce additional heterogeneity due to the vaccine target. However, even with this restriction there was substantial heterogeneity in both the antibody measurements and vaccine effectiveness estimates between studies (**Fig. 1-3**). To link vaccine effectiveness studies with predicted antibody levels, we had to assume that binding and neutralizing antibody levels are strongly correlated^45, 46^, i.e. that the relative differences in both binding and neutralizing antibody levels between people on immunosuppressive treatments and healthy controls agree well. To be able to link Ab and VE data from studies involving different variants and different numbers of doses, we also had to adjust neutralizing Ab titers using antibody data from the general (immunocompetent) population from the Stanford Database^38^. This approach assumes that the fold-difference in neutralization titers between variants, vaccines, and doses from data in the general population agrees well with the fold-difference in antibody binding levels in people on immunosuppressive therapies. There are also several differences between the correlate of protection in the general population we used for comparison^1^ and the relationship between Ab levels and vaccine effectiveness we estimated here in people on immunosuppressive therapies: the former was based on randomized controlled trials rather than non-randomized effectiveness trials and these CoPs were estimated using different methods. In addition, neutralization titers in the CoP for the general population were from the corresponding phase II vaccine trials for each vaccine rather than from a separate study that was linked to VE data using adjustments for different doses and variants.

Despite these limitations on the available evidence, this work highlights the significant impact of different immunosuppressive treatments on vaccine immunogenicity. In particular, it highlights the strong impact of B-cell depletion and S1P inhibitor treatments on Ab levels in contrast to a small reduction in Ab levels in people receiving csDMARDs and integrin inhibitors. It further provides evidence that protection against SARS-CoV-2 infection and hospitalization is lower in people on immunosuppressive therapies than in the general population – even at the same Ab level. Finally, our results support the benefit of vaccination for people on immunosuppressive therapies, while also highlighting the increased risk of SARS-CoV-2 infection and hospitalization.

## Materials and methods

### Search strategy and selection criteria

We focused on the same cohorts of people on immunosuppressive therapies as in our systematic review of COVID-19 vaccine effectiveness (Supplement p. 6, Table S9), i.e. we included healthy/immunocompetent controls and people with hematological malignancies (Haem), mixed immune-mediated inflammatory disorders (IMIDs), neurological disorders (Neur), or rheumatic diseases (Rheu)^16^. We categorized treatments in eight different treatment categories: B-cell depletion, csDMARDs (conventional synthetic Disease Modifying Anti-Rheumatic Drugs), integrin inhibitors, anti-IL6R, JAKi (JAK inhibitors), S1P inhibitors, anti-TNFα, and untreated (Table S2).

We performed a systematic search for studies reporting binding antibody levels post-booster mRNA vaccination (3 doses which was the recommended primary vaccination series for people on immunosuppressive therapies in many countries^47–49^) in cohorts with the abovementioned underlying conditions (Haem, IMIDs, Neur, and Rheu) and immunosuppressive treatments. We searched PubMed, Scopus and Embase for research articles published between November 2019 and 30 June 2023. We included studies reporting binding antibody levels after SARS-CoV-2 vaccination in people on immunosuppressive therapies who received predominantly (>80%) mRNA vaccines, reported binding antibody concentrations or titers after three doses and were published in English. Binding antibody levels for other doses were also extracted if they were reported and in the final data we had more 2-dose data than 3-dose date due to studies reporting Ab levels at multiple time points after the second mRNA vaccination (Table S1). We excluded studies in other patient cohorts (e.g. organ or stem cell transplant recipients, people living with HIV), studies where booster (third dose) vaccines were selectively administered to weak- or non-responders to SARS-CoV-2 vaccines, studies not reporting binding antibody levels in a control cohort (immunocompetent/general population controls; these Ab levels could be reported after any dose/number of vaccinations for which there was also data in people on immunosuppressive therapies available in the same study), studies including >20% of individuals with a previous SARS-CoV-2 infection, cohorts with changes to the immunosuppressive treatment during the study, and studies that were published as reviews, editorials, commentary, letters, or conference abstracts. Details of search terms, inclusion and exclusion criteria, and PRIMSA flow-chart are reported in the Supplement (pp. 3-5, Fig. S1).

### Statistical analysis of antibody data

For statistical analysis of (geometric mean) antibody levels in people on immunosuppressive therapies and controls, we included binding Ab levels for any number of mRNA vaccinations (all extracted data were Ab levels after two or three vaccinations). Ab levels were analyzed on a log_10_-scale (thus Ab levels that were reported as “0” were excluded). All geometric mean Ab levels were included in the data analysis and Ab levels at/below the lower limit of detection or at/above the upper limit of detection were excluded in a sensitivity analysis (see Supplement pp. 9-11). We fitted linear mixed-effects models to the Ab data with outcome geometric mean Ab level (log_10_-transformed), random intercept by study, and weighting by the study size (using the lme4 package)^50^. Hypothesis testing was performed (e.g. to test predictive value of conditions and treatments) using likelihood ratio tests and contrast analysis (‘contest’ function from the lmerTest package^51^, Supplement p. 7). Models were compared using the AICc (Akaike Information Criterion with small sample size corrections).

For all statistical analyses, a significance level of 0.05 was used, i.e. p-value <0.05. All confidence intervals are 95% confidence intervals, and all statistical analyses were performed using R (version 4.5.2)^52^.

### Antibody levels and vaccine effectiveness in people on immunosuppressive therapies

We linked vaccine effectiveness (VE) studies with predicted Ab levels to investigate the VE-antibody-relationship in people on immunosuppressive therapies and compare it with an established relationship between VE and neutralizing antibodies in a general population^1^. VE estimates were from a systematic review of VE in people with the same underlying conditions (Table S9)^16^. To link VE estimates to Ab levels, we used data from Khoury et al.^1^ (neutralizing titers against ancestral SARS-CoV-2 after Pfizer or Moderna vaccination), the Stanford University Coronavirus Antiviral & Resistance Database (Stanford Database, SDB; to adjust for different numbers of vaccinations and circulating variants)^38^, and data from this meta-analysis of Ab levels in people on immunosuppressive therapies (to adjust for different underlying conditions or immunosuppressive treatments) (Supplement pp. 8-9).

To account for immunosuppression, we assumed that the ratio of antibody binding levels between healthy controls and people on immunosuppressive therapies after vaccination is indicative of the ratio of neutralizing antibody titers and applied these ratios to the antibody levels for different vaccines, doses, and variants to arrive at an predicted neutralizing Ab level for each VE estimate (see Fig. S4 for a schematic of linking VE studies with Ab levels).

### Analysis of vaccine effectiveness data with predicted Ab levels

The VE data includes effectiveness estimates of vaccinated people on immunosuppressive therapies compared to unvaccinated people on immunosuppressive therapies and was analyzed (after linking with predicted Ab levels) using a random-effects meta-analysis^16^. The effect measure (outcome) was vaccine effectiveness (VE, in some cases calculated from event data, relative risks, odds ratios, or hazard ratios, which were all treated as equivalent). Vaccine effect measures were aggregated via a random-effects meta-analysis using inverse variance weighting and a random intercept by study (metafor package, using the rma.mv function, with restricted maximum likelihood method; for model comparisons we used the maximum likelihood method instead, as indicated below each supplementary table)^53^. The variance for each reported vaccine effect was calculated from the 95% confidence intervals for VE estimates (or other effect measure). We analyzed outcomes infection, hospitalization, and death separately and included a moderator to allow VE to differ by the log_10_-Ab level. Moderator analysis was conducted with potential additional predictors: study type, vaccine, dose, effect measure (relative risk, odds ratio, or hazard ratio, with or without adjustment), circulating variant group, treatment, and condition (Supplement pp. 7-8, p. 10). We tested the impact of these additional moderators on the meta-analysis by determining if adding them improved model fit (based on AICc).

### Role of the funding source

The funder of the study had no role in study design, data collection, data analysis, data interpretation, or writing of the report.

## Data Availability

All data and code are available upon request to the authors and will be made publicly available upon publication.

## Contributors

SCS, DSK, MPD, DC, and SRK developed the concept for the literature review. SRK performed the systematic searches for vaccine effectiveness studies. SRK, KME, ES, CP, PH, RVH, SCS, and RVJ extracted and verified antibody data. ES performed data analysis. ES, DSK, MPD, SRK, TT, and SCS wrote the manuscript and appendix.

All authors had full access to all the data in the study, revised the manuscript, including panels, figures, and tables, and had final responsibility for the decision to submit for publication.

## Declaration of interests

The authors declare no competing interests.

## Acknowledgments

This work is funded by the National Health and Medical Research Council (NHMRC, Australia) Investigator Grants 2034282 (to ES), 2026360 (to DC), 2034108 (to MPD), 2033318 (to DSK), by the Medical Research Future Fund (MRFF, Australia) MRFF 2016062 (to MPD, DSK) and MRFF 2015313 (to SCS and MPD), and by an Australian Government Research Training Program (RTP) Scholarship (to KME).

## Notes

### Competing Interest Statement

The authors have declared no competing interest.

### Author Declarations

Source data were openly available and were identified in a systematic review of data bases PubMed, Scopus, and Embase. Search terms for each data base, selection criteria, PRISMA flow chart, and details for data extraction are all provided in the supplement. Included studies are listed in Table S1 (including the extracted data used in the meta-analysis) in the supplement and are referenced in both the manuscript and the supplement with DOIs for each of the 19 included studies.

