## Supplement for "Immunogenicity and vaccine effectiveness of COVID-19 vaccines in people on immunosuppressive therapies"

**SYSTEMATIC SEARCH OF BINDING ANTIBODY LEVELS IN PEOPLE ON IMMUNOSUPPRESSIVE THERAPIES 3**
Data sources 3
Search strategies for databases 3
Inclusion and exclusion criteria 4
Study selection 5
Data extraction 5

**VACCINE EFFECTIVENESS IN PEOPLE ON IMMUNOSUPPRESSIVE THERAPIES 6**

**SUPPLEMENTARY METHODS 7**
Binding Ab data in people on immunosuppressive therapies 7
Model comparison and selection by AICc 7
Linking vaccine effectiveness and antibody level data 8
Sensitivity analysis: excluding censored Ab levels 9

**SUPPLEMENTARY RESULTS 10**
Model selection by AICc 10
Sensitivity analysis: excluding censored Ab levels 10

**SUPPLEMENTARY FIGURES 12**
Fig. S1 PRISMA flowchart for binding antibody levels in people on immunosuppressive therapies 12
Fig. S2 Binding antibody data by treatment and condition 13
Fig. S3 Binding Ab levels by treatment, doses, and time since last vaccination 14
Fig. S4 Schematic for predicting antibody levels for corresponding vaccine effectiveness estimates 15
Fig. S5 Vaccine effectiveness by Ab level for outcome death 16
Fig. S6 Ab data by treatment and condition after removing censored Ab levels 17
Fig. S7 Binding Ab data for each treatment and condition after removing censored Ab levels 18
Fig. S8 Vaccine effectiveness by Ab level for all outcomes using uncensored Ab data for people on immunosuppressive therapies 19

**SUPPLEMENTARY TABLES 20**
Table S1. Data sources for binding antibody levels 20
Table S2. Treatment and condition categories in the Ab data 33
Table S3. Model estimates for Ab levels in immunosuppressed compared to immunocompetent 34
Table S4. Impact of treatments and conditions on Ab levels 34
Table S5. Impact of treatments and conditions on Ab levels after removing censored Ab levels 36
Table S6. Ab data – model selection by lowest AICc 38
Table S7. Ratio of Ab levels between treatments and conditions 38
Table S8. Ratio of Ab levels between treatments and conditions after removing censored Ab levels 39

Table S9. Vaccine effectiveness (VE) studies and linking method to Ab data 39
Table S10. Univariate meta-analysis of VE by $log_{10}$-Ab levels 41
Table S11. VE data with Ab levels – model selection by lowest AICc 42
Table S12. VE data with Ab levels – lowest AICc models for outcomes infection and hospitalization 43
Table S13. VE data with Ab levels – lowest AICc models with Ab levels excluding censored data 44

**ABBREVIATIONS 45**

**REFERENCES 47**

### Systematic Search of Binding Antibody Levels in People on Immunosuppressive Therapies

We conducted a systematic search according to the PRISMA updated guideline for reporting systematic reviews[1].

#### Data sources

We searched PubMed, Scopus and Embase to identify eligible articles published from 1 November 2019 till 30 June 2023. We did not search for unpublished studies or other literature.

#### Search strategies for databases

1. PubMed

#1 ("hematologic*"[Title/Abstract] AND "malignanc*"[Title/Abstract]) OR ("haematologic*"[Title/Abstract] AND "malignanc*"[Title/Abstract]) OR ("Leukemia"[Title/Abstract] OR "leukaemia"[Title/Abstract] OR "lymphoma"[Title/Abstract] OR "multiple myeloma"[Title/Abstract]) OR ("immune mediated inflammat*"[Title/Abstract] OR "Rheumatoid arthritis"[Title/Abstract] OR "arthritis"[Title/Abstract] OR "rheumatic"[Title/Abstract] OR "inflammatory bowel disease"[Title/Abstract] OR "multiple sclerosis"[Title/Abstract] OR "MS"[Title/Abstract]) OR "myelofibrosis"[Title/Abstract]

#2 "COVID-19"[Title/Abstract] OR "coronavirus"[Title/Abstract] OR "SARS-CoV-2"[Title/Abstract]

#3 "vaccin"[Supplementary Concept] OR "vaccin"[All Fields] OR "vaccination"[MeSH Terms] OR "vaccination"[All Fields] OR "vaccinable"[All Fields] OR "vaccinal"[All Fields] OR "vaccinate"[All Fields] OR "vaccinated"[All Fields] OR "vaccinates"[All Fields] OR "vaccinating"[All Fields] OR "vaccinations"[All Fields] OR "vaccination s"[All Fields] OR "vaccinator"[All Fields] OR "vaccinators"[All Fields] OR "vaccine s"[All Fields] OR "vaccined"[All Fields] OR "vaccines"[MeSH Terms] OR "vaccines"[All Fields] OR "vaccine"[All Fields] OR "vaccins"[All Fields]

#4 "treat*"[All Fields] OR "therap*"[All Fields]

#5 2019/11/01:2023/06/30[Date - Publication]

#6 #1 AND #2 AND #3 AND #4 AND #5
Filters: English

2. Scopus

#1 ((TITLE-ABS-KEY ("hematologic*" AND "malignanc*")) OR (TITLE-ABS-KEY ("haematologic*" AND "malignanc*")) OR (TITLE-ABS-KEY (leukemia OR leukaemia)) OR (TITLE-ABS-KEY (lymphoma OR "multiple myeloma"))) OR (TITLE-ABS-KEY ("immune mediated inflammat*" OR "rheumatoid arthritis" OR rheumatic OR ("inflammatory bowel disease") OR ("multiple sclerosis" OR ms))) AND (TITLE-ABS-KEY ("COVID-19" OR coronavirus OR "SARS-CoV-

2" )) AND (TITLE-ABS-KEY (treat* OR therap*)) AND (TITLE-ABS-KEY (vaccine)) AND PUBYEAR > 2019 AND PUBYEAR < 2023 AND (LIMIT-TO (LANGUAGE,"English"))

3. Embase
#1 ((hematologic* and malignanc*) or (Haematologic* and malignanc*) or (leukemia or leukaemia) or lymphoma or "multiple myeloma").mp.

#2 ("immune mediated inflammat*" or "rheumatoid arthritis" or arthritis or rheumatic or "inflammatory bowel disease" or " multiple sclerosis" or MS).mp.

#3 (coronavirus or "COVID-19" or "SARS-CoV-2").mp.

#4 (treat* or therap*).mp.

#5 Vaccine/
#6 (#1 OR #2) AND #3 AND #4 AND #5

limit to (english language and yr="2020 - 2023")

Manually removed articles (15) published after June 2023

#### Inclusion and exclusion criteria

Inclusion criteria were: (i) Adults receiving moderate to severe secondary immunosuppression for the treatment of a hematological malignancy (excluding bone marrow transplant) or autoimmune/inflammatory disorder (inclusive of multiple sclerosis and inflammatory bowel disease); (ii) received predominantly (>80%) SARS-CoV-2 mRNA vaccination (only ancestral virus vaccines); (iii) reported post-booster binding antibody (Ab) levels (3 dose levels) for an immunosuppressed cohort; (iv) reported Ab levels for healthy controls (at any number of vaccinations for which the study also reported Ab levels in people on immunosuppressive therapies); (v) published in English.

We excluded:

(i) studies or sub-groups that included
  - patients with stem cell or organ transplant;
  - HIV patients;
  - patients suffering from cancers other than hematological malignancy; or
  - >20% of individuals with a previous SARS-CoV-2 infection;

(ii) cohorts with changes in immunosuppressive treatments during the study, an immunosuppressive treatment type not in the list of included treatment types, or a combination/mixture of treatment types;

(iii) studies where vaccine booster doses were selectively administered to weak- or non-responder subgroup;

(iv) studies with no Ab levels for healthy controls;

(v) reviews, editorials, commentary, letters/correspondence to the editor, conference abstracts.

A full list of included condition categories and treatment types as well as the specific conditions and treatments in the included studies is shown in Table S2.

#### Study selection

Using Covidence (Veritas Health Innovation, Melbourne, Australia), two reviewers (SRK, ES) independently screened the title and abstract of studies identified, excluded studies that were irrelevant and reviewed the full text of all potentially eligible studies. Any disagreements were resolved by discussion.

#### Data extraction

Two reviewers (ES, SRK) extracted data from the included studies. We extracted: first author name, year of publication, country of origin, study design, characteristics of study population including the control group (number of study participants, age, sex, immunosuppressive treatments and condition category), details of the intervention (type and number of vaccines taken, time since last vaccination), information about the antibody assay (assay type and name, SARS-CoV-2 antigen, time of serum collection, limit of detection), and outcome (mean or median binding Ab level with standard deviation, standard error, or 95% confidence interval, and whether the Ab level was below the lower limit of detection or above the upper limit of detection). While we required that studies reported 3-dose Ab levels in people on immunosuppressive therapies, we extracted Ab level data for all reported doses. Extracted data was checked by another author (SRK, KME, ES, CP, PH, RVH, SCS, or RVJ) (see Table S1 for the extracted data).

### Vaccine Effectiveness in People on Immunosuppressive Therapies

We used data for vaccine effectiveness (VE) in people on immunosuppressive therapies from a published systematic review[2].

In short, we searched PubMed, Scopus, and Embase to identify eligible articles published till 5 December 2025. We did not search for unpublished studies or other literature.

Inclusion criteria were: (i) Adults receiving moderate to severe secondary immunosuppression for the treatment of a hematological malignancy (excluding bone marrow or solid organ transplants) or autoimmune/inflammatory disorder (inclusive of multiple sclerosis and inflammatory bowel disease); (ii) received any type of SARS-CoV-2 vaccination; (iii) published in English.

We excluded studies that (i) included patients with stem cell or organ transplant; (ii) HIV patients; (iii) patients suffering from cancers other than hematological malignancy; (iv) reviews, editorials, commentary, letters/correspondence to the editor, conference abstracts.

To match the Ab level data, we included only VE studies in which participants received predominantly (>80%) SARS-CoV-2 mRNA vaccinations (Pfizer, Moderna, or a combination of these vaccines). Included studies are listed in Table S9 with vaccine, doses, variant, conditions, treatments, and information used for linking with Ab data (more details, including vaccine effectiveness for different outcomes, can be found in Table S1 of Stadler et al. (2026))[2].

### Supplementary Methods

#### Binding Ab data in people on immunosuppressive therapies

Studies included in the binding antibody (Ab) data are shown in Table S1 and conditions and treatments were categorized as described in Table S2.
In addition to immunosuppressive treatments and conditions, we included a binary variable indicating immunocompetency ("TRUE" or "FALSE", based on absence or presence of included immunosuppressive conditions, respectively). Vaccines were categorized as Pfizer, Moderna, or mRNA if there was a mixture of vaccines with at least 80% Pfizer or Moderna vaccines (Ab levels after vaccination with >20% non-mRNA vaccines were excluded). Doses administered were 2 or 3 doses with 2 doses as the reference for statistical analysis (the antibody data contains more data for 2 doses than for 3 doses). We excluded baseline Ab levels measured before or on the day of the first vaccination. Time since last vaccination groups are within two months ("<2m", the reference for statistical analysis), between two and six months ("2-6m"), or more than six months (">6m"). If a study included Ab levels measured on the day of a vaccination, we used information on the time between vaccinations to include these Ab levels at the previous dose with the time since last vaccination informed by the time between vaccinations. Ab levels above the upper limit of quantification and below the lower limit of quantification were included in the main analysis but were excluded in a sensitivity analysis (see Sensitivity analyses). Ab levels for patients receiving a combination of the treatment categories listed in Table S2 were excluded to distinguish treatment and condition effects, but Ab levels for patients with immunosuppressive conditions who did not receive immunosuppressive treatment (treatment information included "no therapy", "untreated", "no DMT" (Disease Modifying Treatment)) were included in the analysis.

#### Model comparison and selection by AICc

To compare different fixed effects as predictors of $\log_{10}$-Ab levels, we compared and selected models based on AICc (Akaike Information Criterion with small sample size correction) and using the likelihood ratio test (LRT) that compares nested models (using the function 'anova' with test="LRT"). Interactions between predictors were not included.

In Ab data model selection by AICc, we included the following predictors:

- immunocompetent: TRUE (for healthy controls) or FALSE (people who are immunosuppressed regardless of treatment status)
- Treatment (immunosuppressive treatment): Untreated (reference), B-cell depletion, csDMARDs, anti-IL6R, Integrin inhibitor, JAKi, S1P inhibitors, anti-TNF$\alpha$
- Condition (underlying condition for which people received immunosuppressive treatment): Healthy (reference), Haem, mixed IMIDs, Neur, Rheu
- Vaccine: mRNA, Pfizer, Moderna
- Response (SARS-CoV-2 antigen): Spike, TSP (trimeric spike protein), trimeric S1/S2, S1, RBD, SARS-CoV-2

Due to the known impact of the number of vaccine doses and the time since the last vaccine dose, we a priori included in all models these predictors[3]:

- Doses: 2 or 3 (categorical variable)
- DaysPostLastDoseGroup: '<2m' (reference), '2-6m', or '>6m'

To assess the impact of $\log_{10}$-Ab levels and additional moderators on vaccine effectiveness (VE), we evaluated all possible combinations of additional predictors and selected the model with the lowest

AICc. Interactions between additional predictors were not included (to reduce the number of possible models).

We included the following predictors of vaccine effectiveness for model selection by AICc:
- $\log_{10}$(Ab): $\log_{10}$-Ab level from linking Ab and VE data
- StudyType: Test-negative case-control, Retrospective cohort, Nested matched case-control
- Vaccine: Moderna, mRNA, Pfizer
- Doses: 1, 2, 3, 1-3, 2-3
- VariantGroup: pre-Omicron, pre-Omicron to Omicron BA.1, pre-Omicron to Omicron, Omicron BA.1, Omicron BA.1 to BA.5
- Omicron: pre-Omicron, pre-Omicron to Omicron, Omicron
- Treatment: B-cell depletion, mixed/unknown, "csDMARDs, Integrin inhibitors, JAKi, anti-TNF$\alpha$"
- Condition: Haem, mixed IMIDs, Neur, Rheu
- Effect measure: aOR, RR, aHR, OR

The results of model selections by AICc are reported in the Supplementary results.

#### Linking vaccine effectiveness and antibody level data

A schematic overview of the process of linking vaccine effectiveness (VE) data with antibody (Ab) levels is shown in Fig. S4.

The basis for the linked Ab levels are the neutralizing Ab titers for 2 vaccinations with Pfizer and Moderna vaccines against the ancestral variant from Khoury et al. (2021)[4]: 2.37 and 4.14 fold-convalescent for Pfizer and Moderna, respectively. The titer for 2 vaccinations with any mRNA vaccine is the geometric mean titer of the titers for Pfizer and Moderna: 3.13 fold-convalescent.

The titers were adjusted to allow for different numbers of vaccinations and SARS-CoV-2 variants using data from the Stanford University Coronavirus Antiviral & Resistance Database[5]. Ab levels were extracted from the database in October 2023 for statistical analysis. We excluded prior infections, surrogate neutralization assays, pseudo virus neutralization assays, and data from animal studies. We included Pfizer, Moderna, and a mix of Pfizer and Moderna vaccines (ancestral variant vaccines only, as in the vaccine effectiveness data), Ab levels against ancestral (WT), Alpha, Delta, and Omicron BA.1, BA.2, and BA.5 variants, all vaccine doses included in the vaccine effectiveness data, and only the timepoint 1 month after vaccination.
The statistical model is a linear mixed-effects model (using the lmer function of the lmerTest package[6]) for the $\log_{10}$-Ab level with random intercept by reference and a combination of vaccine, variant, and dose (number of vaccinations) as a fixed effect (concatenated to a single predictor, fitted using "REML").
The outcome of the statistical model is the Ab level for different variant-dose combinations relative to 2 doses and ancestral SARS-CoV-2 for Pfizer, Moderna, and mRNA vaccines (and the variances for these relative Ab levels). We used this with the Khoury et al. (2021)[4] fold-convalescent titers to calculate the fold-convalescent Ab levels of a general population for different doses of Pfizer, Moderna, and mRNA vaccines against different variants.

To account for immunosuppression, we used a statistical analysis of the binding Ab data and either the immunosuppressive treatment or condition in the vaccine effectiveness study.
We used linear mixed-effects models (using the lmer function of the lmerTest package[6]) for the $\log_{10}$-Ab level with random intercept by study and fixed effects and weighting by the study size. The fixed effects for all models were: the dose (number of vaccinations), the time since last vaccination ('<2months', '2-6 months', '>6 months'), and either the treatment or the condition category.

The estimates for the fixed effects of treatment or condition (relative to healthy/untreated controls) were used to adjust the neutralizing Ab titers in a general population from Khoury et al. (2021)[4] and the Stanford University Coronavirus Antiviral & Resistance Database[5].
When vaccines, doses, variants, treatments, or conditions were a combination of different vaccines, doses, etc., we used the geometric mean Ab level for each included vaccine, dose etc., e.g., dose 1-3 relative Ab level is the geometric mean of the relative Ab levels after 1, 2, and 3 doses.

Binding antibody data in people on immunosuppressive therapies is limited to Ab levels after mRNA vaccinations. Thus, six VE studies were not matched with Ab levels due to inclusion of non-mRNA vaccines: Barzegar[7] (participants received a whole inactivated virus vaccine), Copland[8] (mixture of mRNA vaccines and viral vector vaccines), Lazar Neto[9] (mixture of mRNA vaccines and viral vector vaccines), Lee[10] (mixture of mRNA and vector vaccines), Pandit[11] (mixed vaccines including viral vector and inactivated virus vaccines), and Sedighi[12] (mixed vaccines including viral vector and inactivated virus vaccines).

#### Sensitivity analysis: excluding censored Ab levels

As a sensitivity analysis of the binding antibody data, we repeated all analyses with censored Ab levels excluded, i.e. we excluded Ab levels outside of the detectable range of the assay: either at or below the lower limit of quantification or at or above the upper limit of quantification (these Ab levels are indicated by * and #, respectively, in Table S1).

Results of this sensitivity analysis are reported in the Supplementary Results.

### Supplementary Results

#### Model selection by AICc

For Ab binding levels, the best model by AICc included the number of vaccinations (2 or 3, categorical variable), the time since last vaccination (<2 months, 2-6 months, or >6 months), and immunosuppressive treatment (Table S2) as predictors of the $\log_{10}$-Ab level (Table S6). The lowest-AICc model agreed with model 3 in Table S5 and included only the immunosuppressive treatment as a predictor relating to immunosuppression (it did not include the underlying condition).

The best model by AICc for predicting vaccine effectiveness against infection included as predictors: the $\log_{10}$-Ab level, number of doses (vaccinations; categorical variable: 1, 2, 3, 1-3, or 2-3), variant group (pre-Omicron, pre-Omicron to Omicron BA.1, pre-Omicron to Omicron, Omicron BA.1, Omicron BA.1 to BA.5), and study type (test-negative case-control, retrospective cohort, nested matched case-control) (Table S11, Table S12).

For outcome hospitalization, the model with the lowest AICc included as predictors the number of vaccine doses, immunosuppressive treatment, variant (grouped as pre-Omicron, Omicron, or a combination of both), and the method used to measure vaccine effectiveness (effect measure: aOR, RR, aHR, OR; Table S11). However, in a sensitivity analysis that excluded censored antibody levels, the model with the lowest AICc included as predictors the number of doses, effect measure, and $\log_{10}$-Ab level (see below and Table S11).

For outcome death, we did not compare models by AICc due to the low amount of data (5 data points from 3 studies, Fig. S5).

#### Sensitivity analysis: excluding censored Ab levels

Excluding censored Ab levels (i.e. Ab levels outside of the detectable range that were reported as at the lower limit of quantification or at the upper limit of quantification) reduced the number of observations from 169 (all data) to 147 (only detectable Ab levels). The excluded censored Ab levels were from Capuano[13] (n=5 data points for B-cell depletion and S1P inhibitor treatments), Conway[14] (n=2 data points for B-cell depletion and 1 for healthy controls at the upper limit of detection), Furer[15] (n=2 data points for B-cell depletion and 6 at the upper limit of detection for healthy controls and csDMARDs, anti-TNF$\alpha$, anti-IL6R, and JAKi treatments), Palomares Cabeza[16] (n=3 data points for B-cell depletion and S1P inhibitor treatments), Rosati[17] (n=1 for B-cell depletion), and Ruggieri[18] (n=2 data points for B-cell depletion treatment) (Table S1). No Ab study was excluded entirely, and Ab levels were available for the same condition and treatment combinations as before excluding censored Ab levels (Fig. S6).

Ab levels for people on immunosuppressive therapies were 4.03-fold lower compared to healthy controls (95% CI: 2.76 – 5.89) compared to 6.41-fold lower with all Ab levels (95% CI: 4.02-10.22, Table S3). We find again stronger evidence for differences in Ab levels due to the immunosuppressive treatment than due to the underlying condition (Fig. 3B, Fig. S7, Table S5, Table S6). B-cell depletion and S1P inhibitor treatments were most affected by the exclusion of censored Ab levels with the relative Ab levels for both treatments decreasing from 54.5- and 24.8-fold lower Ab levels than in untreated and healthy controls to 19.3- and 23.5-fold lower Ab levels, respectively (Fig. 3B, Fig. S7C-D, Table S8A). B-cell depletion and S1P inhibitor treatments remain the treatments associated with the lowest binding Ab levels. Similarly, binding Ab levels decreased for people with neurological conditions, from 34.4-fold to 14.4-fold lower Ab levels than healthy controls (Fig. S7A-B, Table S8B).

After linking Ab data (without censored Ab levels) and VE data, the relationship between VE and $log_{10}$-Ab levels remains significant and comparable to the previous relationship though with a slightly lower protection at the same Ab level (Fig. S8, Table S10). Ab data for outcome death were only minimally affected by the exclusion of censored Ab levels (Fig. S8C).

Overall, our conclusions are not affected by the exclusion of censored Ab levels.

### Supplementary Figures

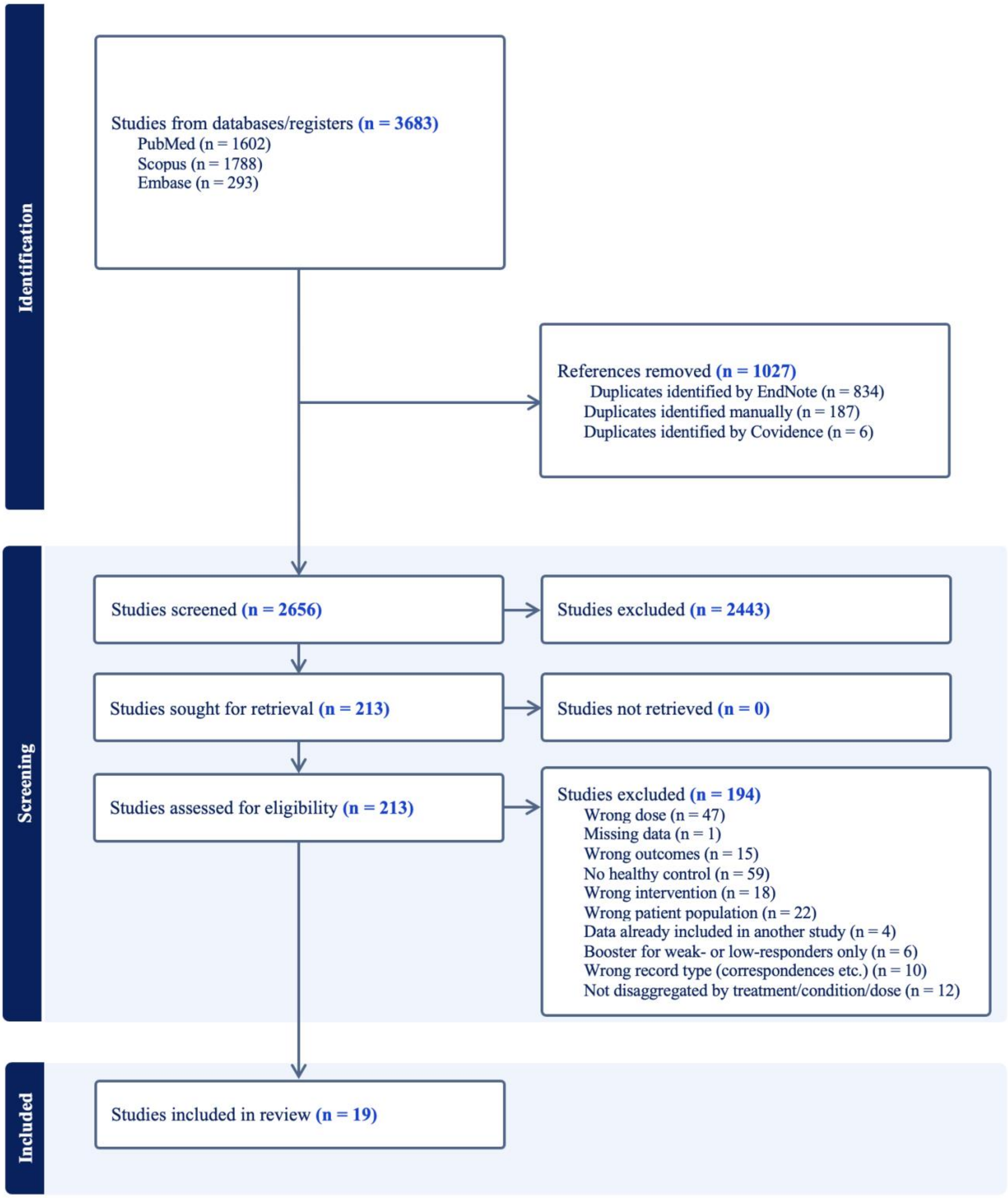


Fig. S1 PRISMA flowchart for binding antibody levels in people on immunosuppressive therapies

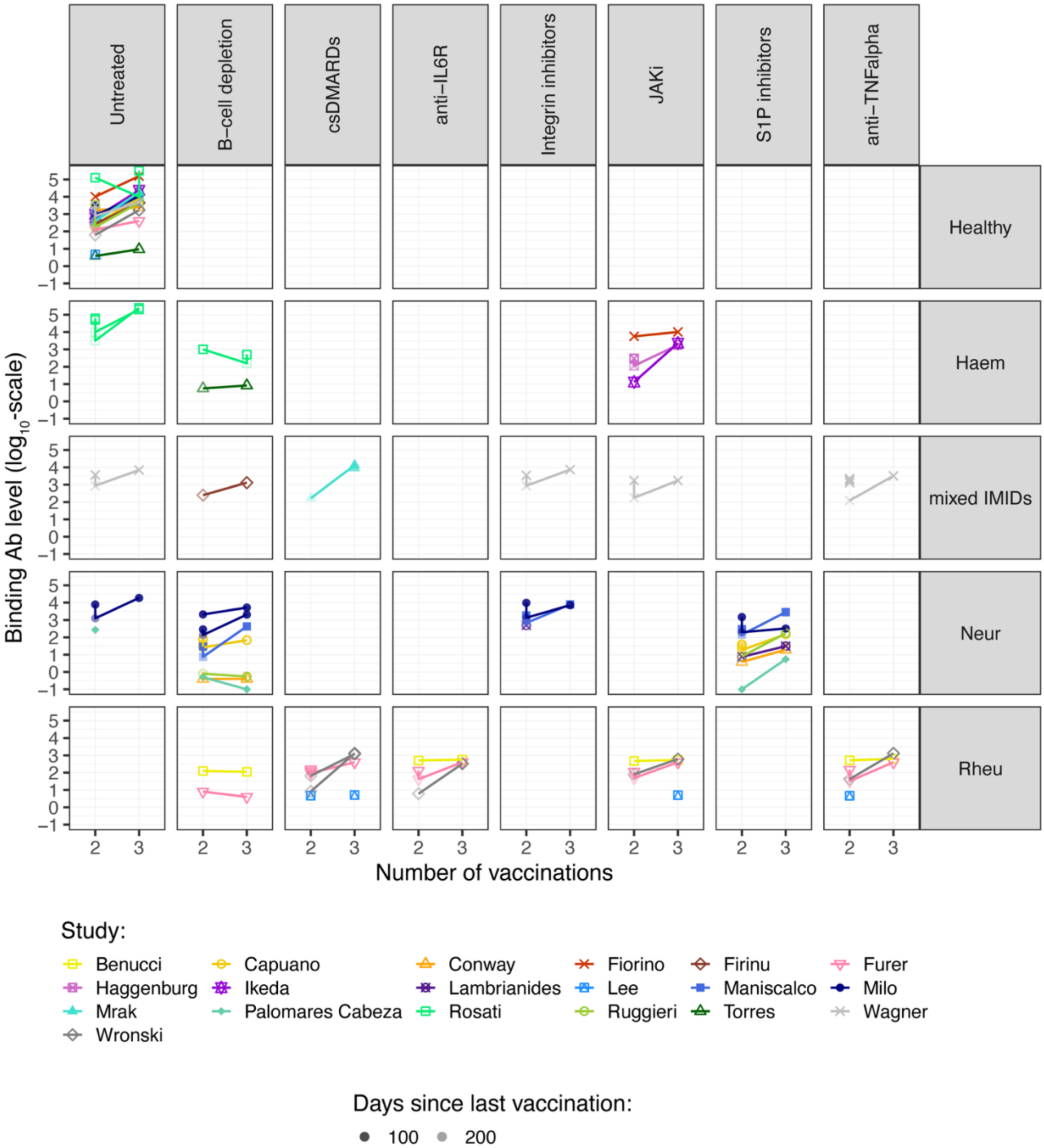


#### Fig. S2 Binding antibody data by treatment and condition

All antibody levels (studies' reported cohort antibody levels, median or geometric mean antibody levels) at any time after vaccination.

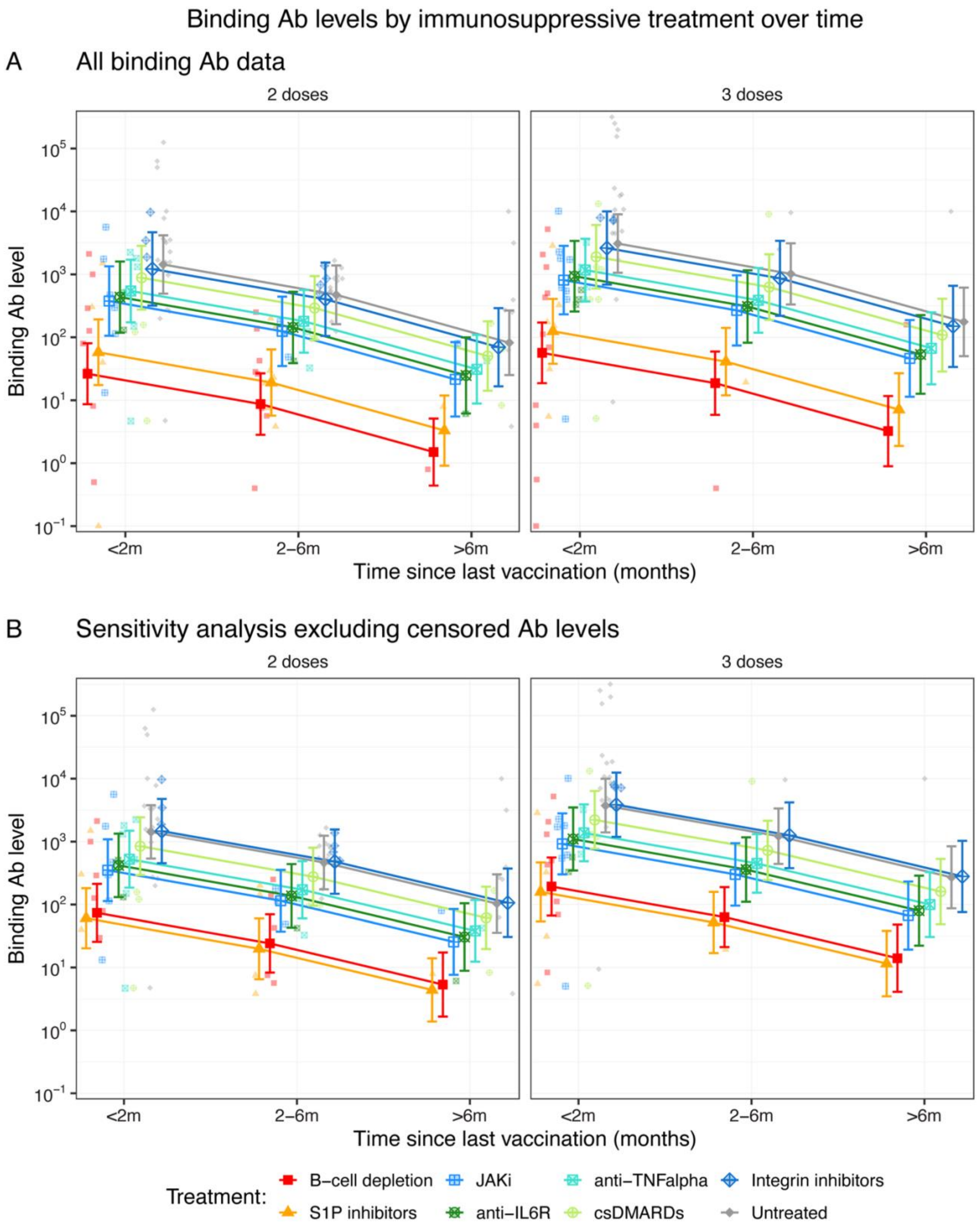


#### Fig. S3 Binding Ab levels by treatment, doses, and time since last vaccination

Visualization of the best (by AICc) statistical model for binding Ab levels which includes the immunosuppressive treatment, number of doses, and time since last vaccination (grouped as "<2 months", "2-6 months", >6 months") (see Table S4-6).

*Neutralizing Ab titers for the general population*

| Vaccine (all 2 doses, ancestral variant) | Titre (fold-convalescent) |
|---|---|
| Pfizer | 2.37 |
| Moderna | 4.14 |
| mRNA | 3.13 |

Source: Khoury et al. (2021).

**Adjust titers to different doses and variants**

*Ab levels for various doses and variants (general population)*

| Vaccine | Doses | Variant | Titer relative to 2 doses, ancestral variant (same vaccine) | Absolute titer (fold-convalescent) |
|---|---|---|---|---|
| Pfizer | 2 | pre-Omicron | 0.678 | 1.609 |
| Moderna | 2 | pre-Omicron | 0.655 | 2.710 |
| mRNA | 1 | pre-Omicron | 0.133 | 0.418 |
| | 2 | | 0.622 | 1.950 |
| | 3 | | 2.266 | 7.101 |
| | 1-3 | | 0.577 | 1.808 |
| | 2-3 | pre-Omicron to Omicron BA.1 | 0.616 | 1.930 |
| | 1 | pre-Omicron to Omicron | 0.060 | 0.188 |
| | 2 | | 0.200 | 0.626 |
| | 3 | | 0.874 | 2.739 |
| | 1-3 | | 0.247 | 0.774 |
| | 2 | Omicron BA.1 | 0.038 | 0.118 |
| | 3 | | 0.470 | 1.471 |
| | 2 | Omicron BA.1 to BA.5 | 0.046 | 0.145 |
| | 3 | | 0.380 | 1.190 |

Source: Stanford University Coronavirus Antiviral & Resistance Database.

**Adjusting for immunosuppression**

**Treatment information available?**

**Yes**

*Relative Ab levels by treatment*

| Treatment | Titers relative to healthy |
|---|---|
| B-cell depletion | 0.018 |
| S1P inhibitor | 0.040 |
| JAKi | 0.262 |
| anti-IL6R | 0.303 |
| anti-TNFα | 0.379 |
| csDMARDs | 0.616 |
| Integrin inhibitors | 0.849 |

Source: This study.

**No**

*Relative Ab levels by condition*

| Condition | Titers relative to healthy |
|---|---|
| Neur | 0.029 |
| Haem | 0.204 |
| Rheu | 0.329 |
| mixed IMIDs | 0.608 |

Source: This study.

#### Fig. S4 Schematic for predicting antibody levels for corresponding vaccine effectiveness estimates

For each vaccine effectiveness estimate after mRNA vaccination that was reported in the literature and identified in Stadler et al. (2026)[2], we predicted a corresponding neutralizing antibody level. The procedure used to predict these levels is depicted here. First, we used the fold-convalescent neutralizing titers from Khoury et al. (2021)[4] which reports 2-dose neutralization titers against the ancestral SARS-CoV-2 variant for mRNA vaccines. We then adjusted these titers for other variants and doses using relative neutralizing Ab levels from an analysis of the Stanford University Antiviral & Resistance Database[5]. Finally, we adjusted for immunosuppression based on either the immunosuppressive treatment or underlying condition using data from our binding Ab level search and meta-analysis. For details see Supplementary Methods, Linking vaccine effectiveness and Ab levels (pp. 8-9). Note this approach assumes that fold changes in the binding antibody levels produced by vaccination is highly correlated with the any reduced neutralization in people on immunosuppressive therapies and that the effect of different numbers of vaccinations (doses) and virus variants is comparable between the general (immunocompetent) population and people on immunosuppressive therapies.

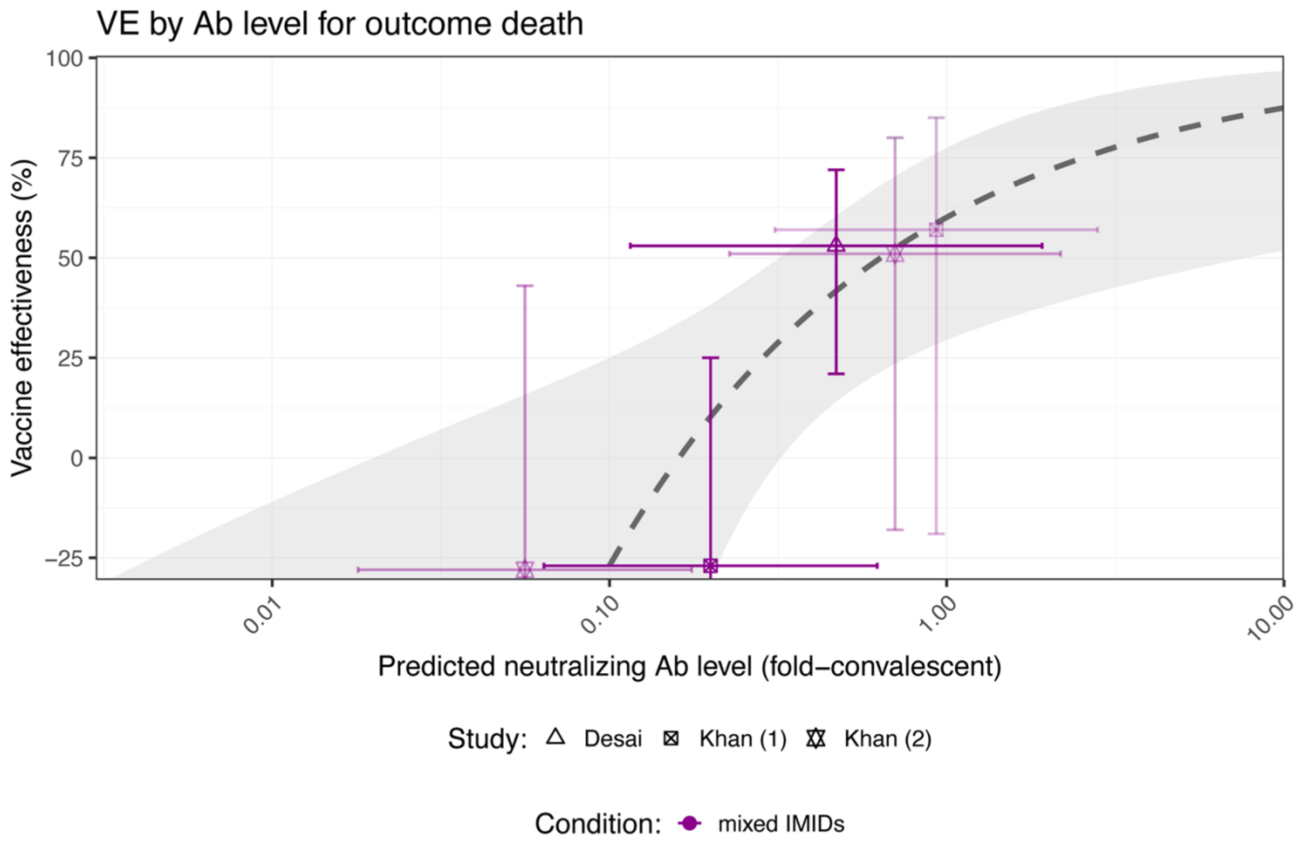


#### Fig. S5 Vaccine effectiveness by Ab level for outcome death

There were only three studies of VE after mRNA vaccination that reported outcome death and there is no established relationship between VE and Ab levels for outcome death in the general (immunocompetent) population for comparison. The fit to the data is the univariate model with $\log_{10}$(Ab) as moderator and a random effect by study.

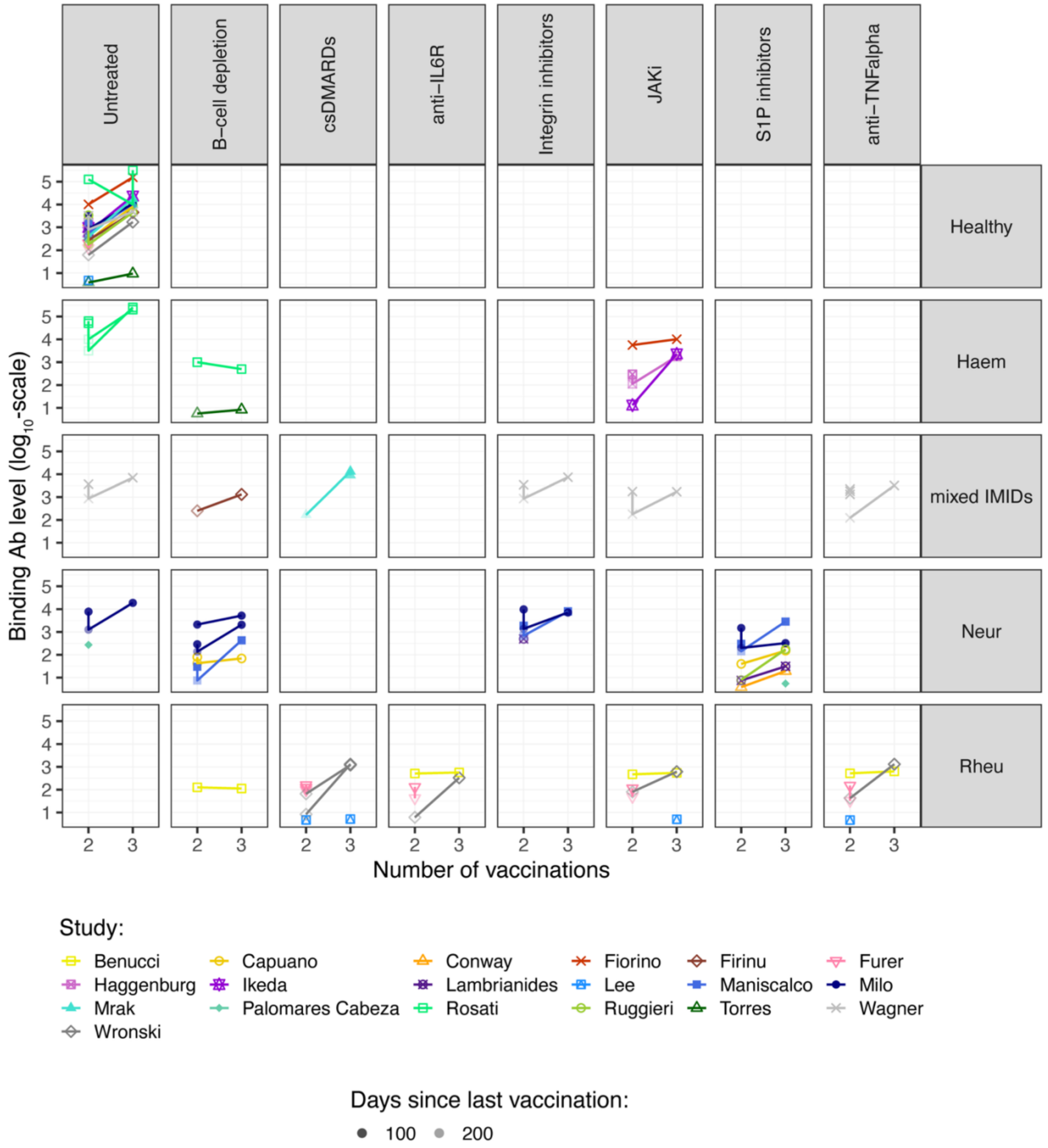


#### Fig. S6 Ab data by treatment and condition after removing censored Ab levels

Data for uncensored Ab levels (i.e. between the lower and upper limit of quantification) and at any time after vaccination.

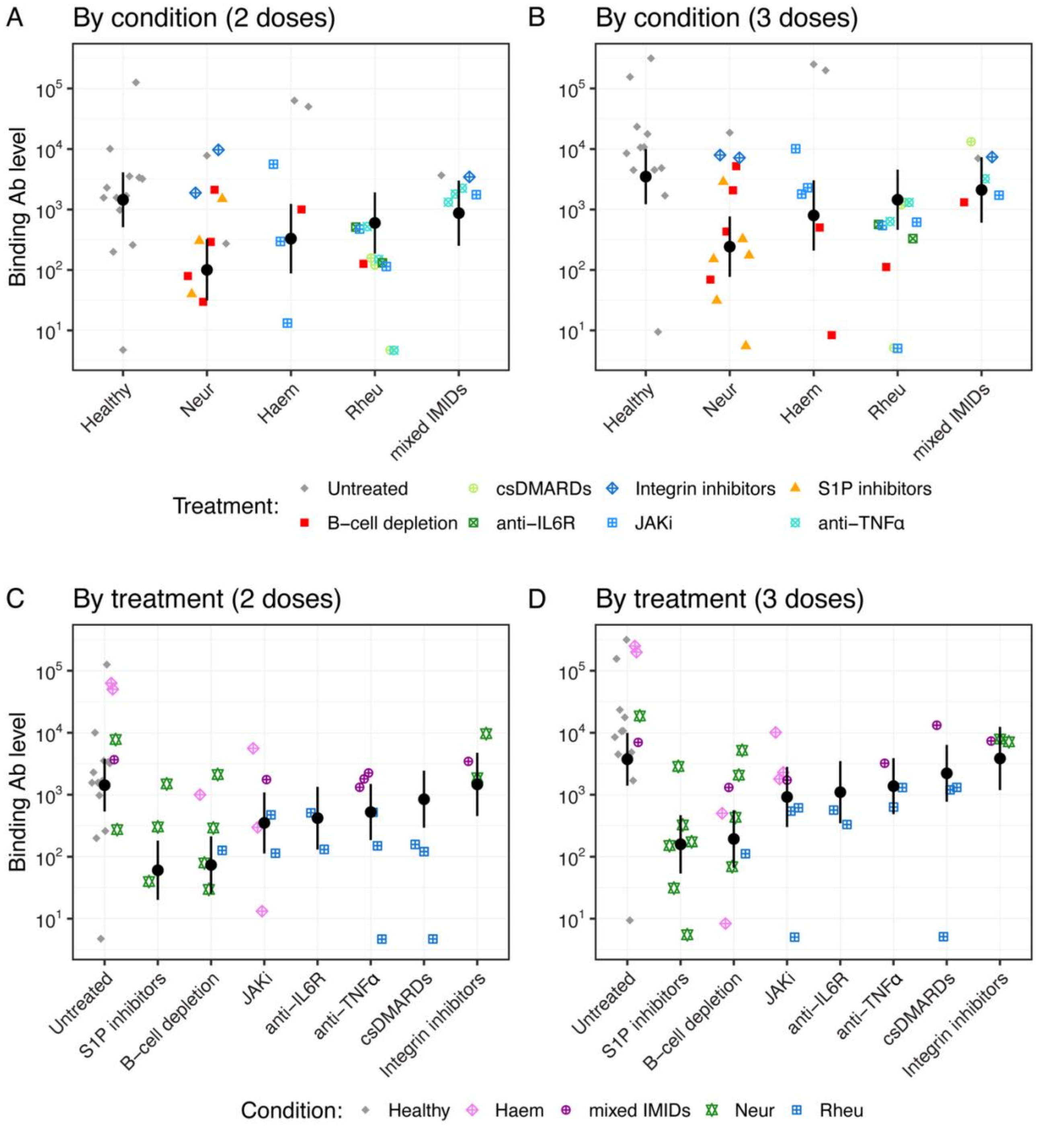


#### Fig. S7 Binding Ab data for each treatment and condition after removing censored Ab levels

Data for uncensored Ab levels (i.e. between the lower and upper limit of quantification) after 2 and 3 doses of an mRNA vaccine and within 2 months after vaccination.

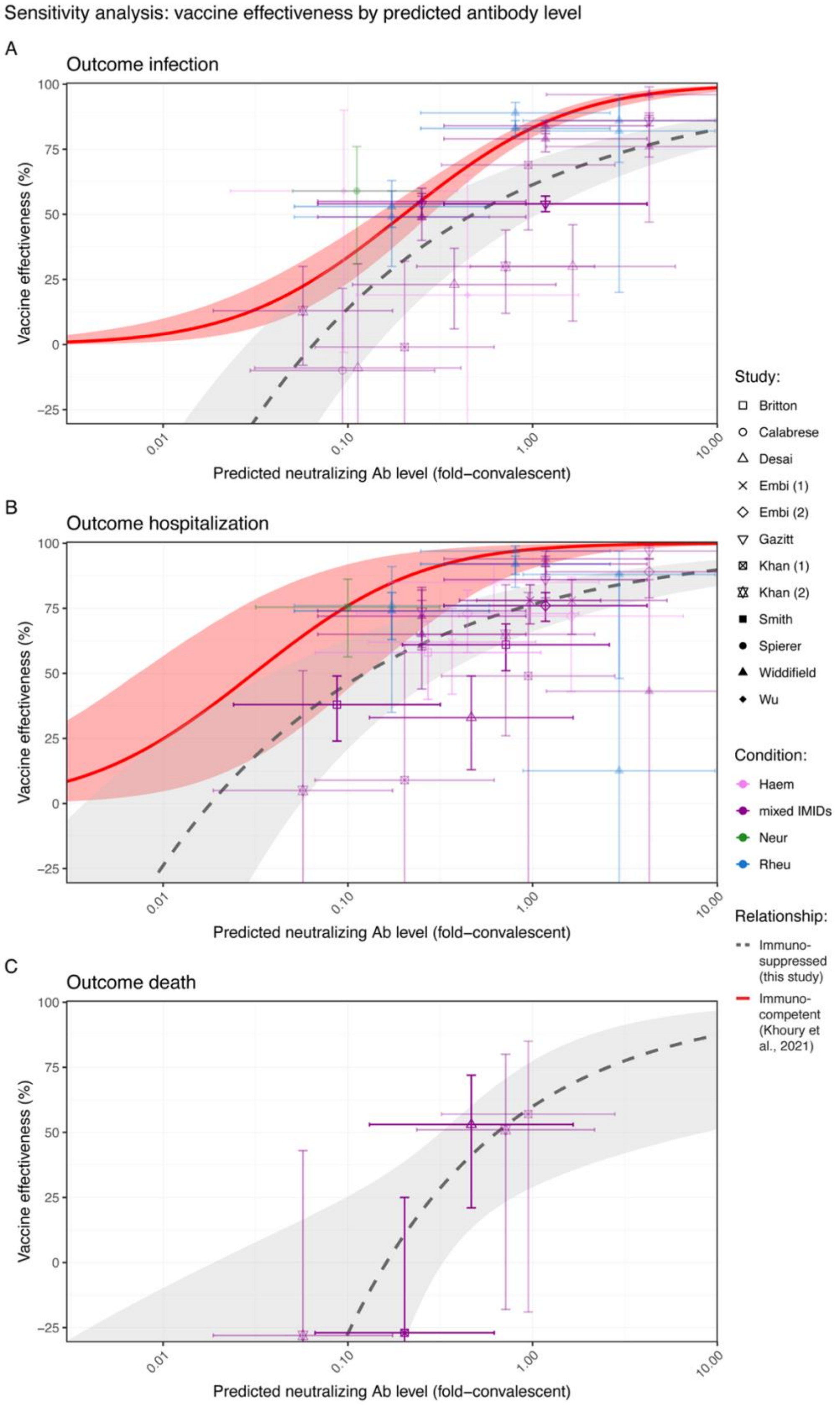


Fig. S8 Vaccine effectiveness by Ab level for all outcomes using uncensored Ab data for people on immunosuppressive therapies

The adjustment for immunosuppression is based on data including only uncensored Ab levels (i.e. between the lower and upper limit of quantification).

### Supplementary Tables

Table S1. Data sources for binding antibody levels

| First author | Study type | Condition (subgroup) | Treatment (drugs) | Vaccine | Doses | Time since last vaccination | Age | Percent female | N | Notes: assay, response, units | Ab level |
|---|---|---|---|---|---|---|---|---|---|---|---|
| Benucci[19] | Prospective | Healthy | Untreated | Pfizer | 2 | <2 months (3 weeks) | 50.54 | | 96 | FEIA, Spike IgG, BAU WHO/mL | 1,562 (IQR: 975-1,632) |
| Benucci[19] | Prospective | Rheu (Rheumatoid arthritis) | anti-IL6R (Sarilumab, Tocilizumab) | Pfizer | 2 | <2 months (3 weeks) | 67.82 | 78 | 64 | FEIA, Spike IgG, BAU WHO/mL | 510 (IQR: 0.70-1,632) |
| Benucci[19] | Prospective | Rheu (Rheumatoid arthritis) | anti-IL6R (Sarilumab, Tocilizumab) | Pfizer | 3 | <2 months (3 weeks) | 67.82 | 78 | 64 | FEIA, Spike IgG, BAU WHO/mL | 565 (IQR: 12-1,632) |
| Benucci[19] | Prospective | Rheu (Rheumatoid arthritis) | JAKi (Baricitinib, Tofacitinib, Upadacitinib) | Pfizer | 2 | <2 months (3 weeks) | 67.82 | 78 | 24 | FEIA, Spike IgG, BAU WHO/mL | 472 (IQR: 218.25-789) |
| Benucci[19] | Prospective | Rheu (Rheumatoid arthritis) | JAKi (Baricitinib, Tofacitinib, Upadacitinib) | Pfizer | 3 | <2 months (3 weeks) | 67.82 | 78 | 24 | FEIA, Spike IgG, BAU WHO/mL | 543 (IQR: 67-1,632) |
| Benucci[19] | Prospective | Rheu (Rheumatoid arthritis) | B-cell depletion (Rituximab) | Pfizer | 2 | <2 months (3 weeks) | 67.82 | 78 | 18 | FEIA, Spike IgG, BAU WHO/mL | 126.5 (IQR: 29.55-568.25) |
| Benucci[19] | Prospective | Rheu (Rheumatoid arthritis) | B-cell depletion (Rituximab) | Pfizer | 3 | <2 months (3 weeks) | 67.82 | 78 | 18 | FEIA, Spike IgG, BAU WHO/mL | 112 (IQR: 9-1,632) |
| Benucci[19] | Prospective | Rheu (Rheumatoid arthritis) | anti-TNFα (Adalimumab, Certolizumab, Etanercept, Golimumab, Infliximab) | Pfizer | 2 | <2 months (3 weeks) | 67.82 | 78 | 43 | FEIA, Spike IgG, BAU WHO/mL | 520 (IQR: 0.7-1,632) |
| Benucci[19] | Prospective | Rheu (Rheumatoid arthritis) | anti-TNFα (Adalimumab, Certolizumab, Etanercept, Golimumab, Infliximab) | Pfizer | 3 | <2 months (3 weeks) | 67.82 | 78 | 43 | FEIA, Spike IgG, BAU WHO/mL | 632 (IQR: 12-1,632) |

| | | | | | | | | | | | |
|---|---|---|---|---|---|---|---|---|---|---|---|
| Capuano[13] | Observational prospective | Healthy | Untreated | Pfizer | 2 | <2 months (5 weeks) | 42.6 | 57.5 | 40 | CLIA, TSP IgG, BAU/mL | 2,290.9 |
| | | | | | 2 | 2-6 months (13 weeks) | | | | | 933.3 |
| | | | | | 2 | 2-6 months (21 weeks) | | | | | 524.8 |
| | | | | | 2 | >6 months (7-9 months) | | | | | 251.2 |
| | | | | | 3 | <2 months (£8 weeks) | | | | | 8,511.4 |
| | | Neur (Multiple sclerosis) | B-cell depletion (Ocrelizumab) | | 2 | <2 months (5 weeks) | 42.3 | 46.4 | 28 | | 79.4 |
| | | | | | 2 | 2-6 months (13 weeks) | | | | | 42.7 |
| | | | | | 2 | 2-6 months (21 weeks) | | | | | 28.2* |
| | | | | | 2 | 2-6 months (~5 months) | | | | | 26.3* |
| | | | | | 3 | <2 months (£8 weeks) | | | | | 69.2 |
| | | | S1P inhibitor (Fingolimod) | | 2 | <2 months (5 weeks) | 45.8 | 42.1 | 19 | | 39.8 |
| | | | | | 2 | 2-6 months (13 weeks) | | | | | 29.5* |
| | | | | | 2 | 2-6 months (21 weeks) | | | | | 22.9* |
| | | | | | 2 | <2 months (~5 months) | | | | | 18.6* |
| | | | | | 3 | <2 months (£8 weeks) | | | | | 151.4 |

| | | | | | | | | | | | |
|---|---|---|---|---|---|---|---|---|---|---|---|
| Conway[14] | Cohort | Healthy | Untreated | mRNA | 2 | 2-6 months (76 days) | 36.6 | 44.8 | 29 | Spike RBD, U/mL | 1,659 |
| | | | | | 3 | 2-6 months (70 days) | 27.8 | 85.7 | 7 | | 2,500[#] |
| | | Neur (Multiple sclerosis) | S1P inhibitor (Fingolimod | | 2 | 2-6 months (95 days) | 51.1 | 71.4 | 7 | | 3.82 |
| | | | | | 3 | 2-6 months (62 days) | 51.8 | 76.2 | 21 | | 19.3 |
| | | | B-cell depletion (Ocrelizumab) | | 2 | 2-6 months (83 days) | 54.9 | 72.7 | 33 | | 0.4* |
| | | | | | 3 | 2-6 months (62 days) | 54.5 | 72.7 | 22 | | 0.4* |
| Fiorino[20] | Prospective | Healthy | Untreated | mRNA | 2 | <2 months (7-30 days) | 52.2 | 45 | 20 | ELISA, Spike IgG, titer | 9,891 |
| | | | | | 3 | <2 months (15 days) | | | | | 147,661 |
| | | Haem (Myelofibrosis) | JAKi (Ruxolitinib) | | 2 | <2 months (7-30 days) | 72 | 75 | 8 | | 3,948 |
| | | | | | 3 | <2 months (15 days) | | | | | 10,240 |
| Firinu[21] | Prospective | Healthy | Untreated | Pfizer | 2 | 2-6 months (~5 months) | 51.5 | 50 | 18 | CLIA, Spike IgG, BAU/mL | 270 (IQR: 107.8-407.8) |
| | | | | | 3 | <2 months (3-4 weeks) | | | | | 4,490 (IQR: 2,080-8,790) |
| | | mixed IMIDs | B-cell depletion (Rituximab, Ocrelizumab) | | 2 | >6 months (~9 months) | 46 | | | | 251 (IQR: 34.4-421.6) |
| | | | | | 3 | <2 months (3-4 weeks) | | | | | 1,310 (IQR: 261.3-7,300) |

| | | | | | | | | | | | |
|---|---|---|---|---|---|---|---|---|---|---|---|
| Furer[15] | Longitudinal observational | Healthy | Untreated | Pfizer | 2 | <2 months (2-6 weeks) | 50.5 | 64.8 | 122 | LIAISON, trimeric Spike S1/S2 IgG, BAU/mL | 199 (R: 45.1-400) |
| | | | | | 2 | 2-6 months (6 months) | | | 116 | | 124 (R: 6.2-400) |
| | | | | | 3 | <2 months (2-6 weeks) | | | 45 | | 400# (R: 308-400) |
| | | Rheu (RA, PsA, SLE, AxSpA, AAV, CTD, LVV, other vasculitis) | csDMARDs | | 2 | <2 months (2-6 weeks) | 58 | 70.2 | 130 | | 157.5 (R: 0-400) |
| | | | | | 2 | 2-6 months (6 months) | | | 109 | | 95 (R: 4.17-400) |
| | | | | | 3 | <2 months (2-6 weeks) | | | 19 | | 400# (R: 219-400) |
| | | | csDMARDs (Methotrexate) | | 2 | <2 months (2-6 weeks) | | | 41 | | 121 (R: 0-400) |
| | | | | | 2 | 2-6 months (6 months) | | | 32 | | 96.75 (R: 5.51-400) |
| | | | | | 3 | <2 months (2-6 weeks) | | | 6 | | 400# (R: 219-400) |
| | | | anti-TNFα | | 2 | <2 months (2-6 weeks) | | | 179 | | 150 (R: 0-400) |
| | | | | | 2 | 2-6 months (6 months) | | | 159 | | 32.6 (R: 0-330) |
| | | | | | 3 | <2 months (2-6 weeks) | | | 22 | | 400# (R: 0-400) |
| | | | anti-IL6R | | 2 | <2 months (2-6 weeks) | | | 39 | | 131 (R: 27.9-400) |
| | | | | | 2 | 2-6 months (6 months) | | | 33 | | 42.2 (R: 0-400) |
| | | | | | 3 | <2 months (2-6 weeks) | | | 13 | | 400# (R: 92.6-400) |

| | | | | | | | | | | | |
|---|---|---|---|---|---|---|---|---|---|---|---|
| | | | B-cell depletion (Rituximab) | | 2 | <2 months (2-6 weeks) | | | 110 | | 8.09* (R: 0-366) |
| | | | | | 2 | 2-6 months (6 months) | | | 89 | | 0† (R: 0-393) |
| | | | | | 3 | <2 months (2-6 weeks) | | | 47 | | 3.97* (R: 0-400) |
| | | | JAKi | | 2 | <2 months (2-6 weeks) | | | 22 | | 114 (R: 0-324) |
| | | | | | 2 | 2-6 months (6 months) | | | 22 | | 48.65 (R: 0-370) |
| | | | | | 3 | <2 months (2-6 weeks) | | | 8 | | 400# (R: 106-400) |
| Haggenburg[22] | Prospective observational cohort | Healthy | Untreated | Moderna | 2 | <2 months (4 weeks) | 57 | 68 | 37 | S1 IgG, BAU/mL | 1,566.5 |
| | | Haem (MPD) | JAKi (Ruxolitinib) | | 2 | <2 months (4 weeks) | 58 | 41.9 | 31 | | 296.2 |
| | | | | | 2 | 2-6 months (5 months) | | | | | 112.2 |
| | | | | | 3 | <2 months (4 weeks) | | | | | 1,795.9 |
| Ikeda[23] | Longitudinal cohort | Healthy | Untreated | mRNA | 2 | <2 months (2-12 weeks) | 35 | 75 | 80 | Spike RBD, U/mL | 975.5 |
| | | | | | 2 | 2-6 months (5 months) | 35 | 75 | 80 | | 619.5 |
| | | | | | 3 | <2 months (25 days) | 35 | 75 | 80 | | 23,371.5 |
| | | Haem (Lymphoid neoplasms, plasma cell dyscrasias, myeloid neoplasia) | JAKi (JAK1/2i) | | 2 | <2 months (2-12 weeks) | 73 | 46.3 | 12 | | 13.2 |
| | | | | | 3 | <2 months (25 days) | 73 | 46.3 | 12 | | 2286.5 (IQR: 637.7-4,670.2) |

| | | | | | | | | | | | |
|---|---|---|---|---|---|---|---|---|---|---|---|
| Lambrianides[24] | Cohort | Healthy | Untreated | Pfizer | 2 | 2-6 months (3 months) | 46.7 | 59.6 | 52 | CMIA, Anti-S1RBD IgG, BAU/mL | 415.6 (IQR: 244.9-686.5) |
| | | Neur (Multiple sclerosis) | Integrin inhibitor (Natalizumab) | | 2 | 2-6 months (3 months) | 45.1 | 75.4 | 26 | | 495.3 (IQR: 199.1-999.5) |
| | | | S1P inhibitor (Fingolimod) | | 2 | 2-6 months (3 months) | 45.1 | 75.4 | 34 | | 7.5 (IQR: 1.8-21.6) |
| | | | | mRNA | 3 | <2 months (2 weeks) | 45.1 | 75.4 | 23 | | 31.15 (IQR: 12.0-91.0) |
| Lee[25] | Cohort | Healthy | Untreated | mRNA | 2 | <2 months (30 days) | 41 | 64 | 76 | ELISA, Anti-S IgG, $log_{10}$-AUC | 4.74 (IQR: 4.67-4.82) |
| | | Rheu (Inflammatory arthritis) | csDMARDs (Methotrexate) | | 2 | <2 months (30 days) | 53 | 65 | 10 | | 4.7 (IQR: 4.37-5.01) |
| | | | anti-TNFα | | 2 | <2 months (30 days) | 53 | 65 | 12 | | 4.67 (IQR: 4.31-5.03) |
| | | | csDMARDs | | 3 | <2 months (30 days) | 62 | 78 | 28 | | 5.13 (IQR: 4.98-5.28) |
| | | | JAKi | | 3 | <2 months (30 days) | 62 | 78 | 17 | | 5.03 (IQR: 4.83-5.24) |
| Maniscalco[26] | Prospective | Healthy | Untreated | Pfizer | 2 | <2 months (21 days) | 41 | 55.6 | 9 | ELISA, Spike IgG, U/mL | 1,684 (SEM: 527.8) |
| | | | | | | 2-6 months (6 months) | | | | | 548.5 (SEM: 133.2) |
| | | | | | 3 | <2 months (21 days) | | | | | 10,618 (SEM: 737.7) |

| | | | | | | | | | | | |
|---|---|---|---|---|---|---|---|---|---|---|---|
| | | Neur (Multiple sclerosis) | Integrin inhibitor (Natalizumab) | | 2 | <2 months (21 days) | 40 | 55.4 | 13 | | 1,881 (SEM: 731.1) |
| | | | | | | 2-6 months (6 months) | | | | | 683.5 (SEM: 141.9) |
| | | | | | 3 | <2 months (21 days) | | | | | 7,936 (SEM: 1,066) |
| | | | S1P inhibitor (Fingolimod) | | 2 | <2 months (21 days) | | | 9 | | 302.1 (SEM: 122.3) |
| | | | | | | 2-6 months (6 months) | | | | | 143.7 (SEM: 63.33) |
| | | | | | 3 | <2 months (21 days) | | | | | 2,865 (SEM: 928) |
| | | | B-cell depletion (Ocrelizumab) | | 2 | <2 months (21 days) | | | 8 | | 29.7 |
| | | | | | | 2-6 months (6 months) | | | | | 7.5 |
| | | | | | 3 | <2 months (21 days) | | | | | 431 (SEM: 246.2) |
| Milo[27] | Prospective cohort | Healthy | Untreated | Pfizer | 2 | <2 months (~7 weeks) | 50 | 53 | 53 | Spike RBD IgG, AU/mL | 3,537 (R: 42-27876) |
| | | | | | 2 | 2-6 months (25 weeks) | | | 16 | | 855 (R: 108-5,790) |
| | | | | | 3 | <2 months (~7 weeks) | | | 21 | | 10,765 (R: 787-33,761) |
| | | Neur (Multiple sclerosis) | Untreated | | 2 | <2 months (~7 weeks) | 46 | 65 | 28 | | 7,778 (R: 770-32,497) |

| | | | | | | | | | | | |
|---|---|---|---|---|---|---|---|---|---|---|---|
| | | | | | 2 | 2-6 months (25 weeks) | | | 17 | | 1,269 (R:120-12,046) |
| | | | | | 3 | <2 months (~7 weeks) | | | 14 | | 18,640 (R: 454-40,000) |
| | | | B-cell depletion (Ocrelizumab) | | 2 | <2 months (~7 weeks) | | | 89 | | 290 (R: 0-7,722) |
| | | | | | 2 | 2-6 months (25 weeks) | | | 38 | | 137 (R: 0-992) |
| | | | | | 3 | <2 months (~7 weeks) | | | 43 | | 2,072 (R: 0-36,108) |
| | | | B-cell depletion (Ofatumumab) | | 2 | <2 months (~7 weeks) | | | 4 | | 2,120 (R: 120-5,826) |
| | | | | | 3 | <2 months (~7 weeks) | | | 2 | | 5,210 (R: 189-10,230) |
| | | | Integrin inhibitor (Natalizumab) | | 2 | <2 months (~7 weeks) | | | 19 | | 9,698 (R: 161-40,000) |
| | | | | | 2 | 2-6 months (25 weeks) | | | 12 | | 1,372 (R: 259-7,258) |
| | | | | | 3 | <2 months (~7 weeks) | | | 14 | | 7,149 (R: 792-33,990) |
| | | | S1P inhibitor (Fingolimod) | | 2 | <2 months (~7 weeks) | | | 37 | | 1,494 (R: 0-19,543) |
| | | | | | 2 | 2-6 months (25 weeks) | | | 13 | | 200 (R: 0-1,856) |
| | | | | | 3 | <2 months (~7 weeks) | | | 24 | | 325 (R: 0-4,586) |

| | | | | | | | | | | | |
|---|---|---|---|---|---|---|---|---|---|---|---|
| Mrak[28] | Prospective | Healthy | Untreated | mRNA | 2 | >6 months (182 days) | 48.9 | 52.4 | 42 | ECLIA, Spike RBD, BAU/mL | 394.1 (IQR: 217.6-640.6) |
| | | | | | 3 | <2 months (4 weeks) | 48.9 | 52.4 | 42 | | 17,697.5 (IQR: 12,529.5-31,889.1) |
| | | | | | 3 | 2-6 months (12 weeks) | 48.9 | 52.4 | 42 | | 9,585.2 (IQR: 6419.0-15,526.0) |
| | | mixed IMIDs | csDMARDs | | 2 | >6 months (238 days) | 57.9 | 78 | 50 | | 166.7 (IQR: 91.2-309.7) |
| | | | | | 3 | <2 months (4 weeks) | 57.9 | 78 | 50 | | 13,215.5 (IQR: 10,543.9-21,078.0) |
| | | | | | 3 | 2-6 months (12 weeks) | 57.9 | 78 | 50 | | 9,062.2 (IQR: 5,441.3-13,708.1) |
| Palomares Cabeza[16] | Prospective | Healthy | Untreated | Moderna | 2 | <2 months (7-10 days) | 42 | 67 | 12 | ELISpot, RBD IgG, AU/mL | 259 (IQR: 240-547) |
| | | Neur (Multiple sclerosis) | Untreated | | 2 | <2 months (7-10 days) | 53 | 70 | 10 | | 273 (IQR: 248-412) |
| | | | B-cell depletion (Ocrelizumab) | | 2 | <2 months (7-10 days) | 44 | 70 | 24 | | 0.5* (IQR: 0.1-1.3) |
| | | | | | 3 | <2 months (7-10 days) | 44 | 70 | 8 | | 0.1* (IQR: 0.1-6) |
| | | | S1P inhibitor (Fingolimod) | mRNA | 2 | <2 months (28 days) | 45 | 67 | 12 | | 0.1* (IQR: 0.1-2.1) |

| | | | | | | | | | | | |
|---|---|---|---|---|---|---|---|---|---|---|---|
| | | | | | 3 | <2 months (7-10 days) | 45 | 67 | 9 | | 5.5 (IQR: 1.5-7.6) |
| Rosati[17] | Prospective | Healthy | Untreated | Pfizer | 2 | <2 months (1 month) | 54 | 64.9 | 34 | ELISA, trimeric Spike and Spike-RBD, titer | 125,892.5 |
| | | | | | 2 | >6 months (8 months) | 54 | | 37 | | 10,000 |
| | | | | | 3 | <2 months (1 month) | 54 | | 37 | | 316,227.8 |
| | | Haem (Waldenstrom's macroglobulinemia) | Untreated | | 2 | <2 months (1 month) | 82 | 66.7 | 9 | | 63,095.7 |
| | | | | | 2 | >6 months (8 months) | 82 | | 8 | | 3,162.3 |
| | | | | | 3 | <2 months (1 month) | 82 | | 8 | | 251,188.6 |
| | | | B-cell depletion (Rituximab, BTKi) | | 2 | <2 months (1 month) | 70 | 63.7 | 10 | | 1,000 |
| | | | | | 2 | >6 months (8 months) | 70 | | 10 | | 158.5* |
| | | | | | 3 | <2 months (1 month) | 70 | | 10 | | 501.2 |
| | | Haem (Multiple myeloma) | Untreated | | 2 | <2 months (1 month) | 69.5 | 35 | 20 | | 50,118.7 |
| | | | | | 2 | >6 months (8 months) | 69.5 | | 19 | | 10,000 |
| | | | | | 3 | <2 months (1 month) | 69.5 | | 19 | | 199,526.2 |
| Ruggieri[18] | Longitudinal prospective | Healthy | Untreated | mRNA | 2 | <2 months (21 days) | 43 | 72.7 | 99 | Anti-RBD IgG, BAU/mL | 3,377 (IQR: 1,647-4,839) |
| | | | | | 2 | 2-6 months (160 days) | 43 | 72.7 | 99 | | 177 (IQR: 95.5-262.6) |
| | | | | | 3 | <2 months (35 days) | 43 | 72.7 | 99 | | 4,516 (IQR: 3,098-5477) |

| | | | | | | | | | | | |
|---|---|---|---|---|---|---|---|---|---|---|---|
| | | Neur (Multiple sclerosis) | B-cell depletion (Ocrelizumab) | | 2 | >6 months (182 days) | 47 | 70.2 | 29 | | 0.8* (IQR: 0.15-6.25) |
| | | | | | 3 | <2 months (35 days) | 47 | 70.2 | 29 | | 0.55* (IQR: 0.27-6.27) |
| | | | S1P inhibitor (Fingolimod) | | 2 | >6 months (182 days) | 47 | 70.2 | 31 | | 7.9 (IQR: 2.7-39.7) |
| | | | | | 3 | <2 months (35 days) | 47 | 70.2 | 31 | | 174 (IQR: 26-514) |
| Torres[29] | Observational | Healthy | Untreated | Pfizer | 2 | >6 months (189 days) | 77 | 80 | 15 | ELISA, Spike IgG, titer | 3.83 |
| | | | | | 3 | <2 months (29 days) | 77 | 80 | 15 | | 9.41 |
| | | Haem (NHL, AIHA, IT/SLE) | B-cell depletion (Rituximab) | Moderna | 2 | 2-6 months (124.5 days) | 61 | 66.7 | 18 | | 5.65 |
| | | | | | 3 | <2 months (28 days) | 61 | 66.7 | 18 | | 8.34 |
| Wagner[30] | Prospective | Healthy | Untreated | mRNA | 2 | <2 months (4 weeks) | 46.1 | 50 | 66 | ELISA, S1 IgG, BAU/mL | 3,206 |
| | | | | | 2 | 2-6 months (5-6 months) | 46.1 | 50 | 66 | | 788.4 |
| | | | | | 3 | <2 months (4 weeks) | 46.1 | 50 | 66 | | 4842.6 |
| | | mixed IMIDs (IBD) | Untreated | | 2 | <2 months (4 weeks) | 44 | 46.9 | 21 | | 3,676 (IQR: 2,926.5-4,758.0) |
| | | | | | 2 | 2-6 months (5-6 months) | 44 | 46.9 | 21 | | 863.7 (IQR: 678.5-1,065.5) |
| | | | | | 3 | <2 months (4 weeks) | 44 | 46.9 | 21 | | 7,004. (IQR: 6,107.4-7,686.4) |

| | | | | | | | | | | | |
|---|---|---|---|---|---|---|---|---|---|---|---|
| | | | anti-TNFα (Infliximab) | | 2 | <2 months (4 weeks) | 44 | 46.9 | 16 | | 1,319 |
| | | | anti-TNFα (Adalimumab) | | 2 | <2 months (4 weeks) | 44 | 46.9 | 38 | | 1,799 |
| | | | anti-TNFα (Golimumab) | | 2 | <2 months (4 weeks) | 44 | 46.9 | 5 | | 2,246 |
| | | | anti-TNFα | | 2 | 2-6 months (5-6 months) | 44 | 46.9 | 59 | | 123.3 |
| | | | anti-TNFα | | 3 | <2 months (4 weeks) | 44 | 46.9 | 59 | | 3,220.9 |
| | | | Integrin inhibitor (Vedolizumab) | | 2 | <2 months (4 weeks) | 44 | 46.9 | 19 | | 3,454 |
| | | | | | 2 | 2-6 months (5-6 months) | 44 | 46.9 | 19 | | 860.1 |
| | | | | | 3 | <2 months (4 weeks) | 44 | 46.9 | 19 | | 7,361.2 (IQR: 6,202.0-8,389.0) |
| | | | JAKi | | 2 | <2 months (4 weeks) | 44 | 46.9 | 2 | | 1,754.8 |
| | | | | | 2 | 2-6 months (5-6 months) | 44 | 46.9 | 2 | | 178.4 |
| | | | | | 3 | <2 months (4 weeks) | 44 | 46.9 | 2 | | 1,720.4 (IQR: 1,362.3-2,010.5) |
| Wronski[31] | Prospective cohort | Healthy | Untreated | mRNA (Pfizer, Moderna, AstraZeneca, J&J, >80% Pfizer or Moderna) | 2 | >6 months | 48.6 | 71.7 | 43 | ELISA, IgG, RU/mL | 62.2 (R: 8.2-1326.9) |
| | | | | | 3 | <2 months (4 weeks) | 48.6 | 71.7 | 47 | | 1,693.1 (R: 530.5-5,419.1) |
| | | Rheu (Inflammatory arthritis) | csDMARDs (Methotrexate) | | 2 | >6 months | 53 | 68 | 21 | | 65.7 (R: 2.1-303.9) |

| | | | | | | | | | | | |
|---|---|---|---|---|---|---|---|---|---|---|---|
| | | | | | 3 | <2 months (4 weeks) | 53 | 68 | 24 | | 1,199.4 (R: 30.8-4,724.7) |
| | | | csDMARDs (Sulfasalazine) | | 2 | >6 months | 53 | 68 | 5 | | 8.3 (R: 3.4-200.1) |
| | | | | | 3 | <2 months (4 weeks) | 53 | 68 | 6 | | 1,311.2 (R: 30.8-3,950.7) |
| | | | anti-TNFα | | 2 | >6 months | 53 | 68 | 15 | | 42.4 (R: 3.9-637.9) |
| | | | | | 3 | <2 months (4 weeks) | 53 | 68 | 17 | | 1,304.5 (R: 141.6-2,142.2) |
| | | | anti-IL6R | | 2 | >6 months | 53 | 68 | 5 | | 6.1 (R: 1.1-145) |
| | | | | | 3 | <2 months (4 weeks) | 53 | 68 | 5 | | 329.8 (R: 272.2-999.1) |
| | | | JAKi | | 2 | >6 months | 53 | 68 | 5 | | 80 (R: 0-953.8) |
| | | | | | 3 | <2 months (4 weeks) | 53 | 68 | 7 | | 614.8 (R: 95.5-1,870.9) |

* Ab level at or below the lower limit of quantification
### Ab level at the upper limit of quantification
† Ab level 0 data points were excluded from the analysis because Ab levels were analyzed on a $log_{10}$-scale.
Abbreviations: a list of all abbreviation used in the supplement and manuscript is provided at the end of the supplement.

Table S2. Treatment and condition categories in the Ab data

| Treatments | Conditions |
|---|---|
| - Untreated<br><br>- anti-IL6R:<br>• Sarilumab<br>• Tocilizumab<br>- anti-TNFα:<br>• Adalimumab<br>• Certolizumab<br>• Etanercept<br>• Golimumab<br>• Infliximab<br>- B-cell depletion:<br>• BTKi<br>• Ocrelizumab<br>• Ofatumumab<br>• Rituximab<br>- csDMARDs (conventional synthetic Disease Modifying Anti-Rheumatic Drugs)<br>• Methotrexate<br>• Sulfasalazine<br>- Integrin inhibitors:<br>• Natalizumab<br>• Vedolizumab<br>- JAKi:<br>• Baricitinib<br>• JAK1/2i<br>• Ruxolitinib<br>• Tofacitinib<br>• Upadacitinib<br>- S1P inhibitors:<br>• Fingolimod | - Healthy/immunocompetent*<br><br>- Haem: haematological malignancies<br>• Lymphoid neoplasms, plasma cell dyscrasias, myeloid neoplasia<br>• Multiple myeloma<br>• Myelofibrosis<br>• Myeloproliferative disease (MPD)<br>• NHL, AIHA, IT/SLE<br>• Waldenstrom's macroglobulinemia<br>- mixed IMID (immune-mediated inflammatory disorders):<br>• Inflammatory Bowel Disease (IBD)<br>• Not further specified IMIDs<br>- Neur: neurological disorders<br>• Multiple sclerosis<br>- Rheu: rheumatic diseases<br>• Inflammatory arthritis<br>• RA, PsA, SLE, AxSpA, AAV, CTD, LVV, other vasculitis<br>• Rheumatoid arthritis |

* Control individuals from the general population were assumed to be healthy and immunocompetent.
All studies with their treatments and conditions are listed in Table S1.
Abbreviations: a list of all abbreviation used in the supplement and manuscript is provided at the end of the supplement.

#### Table S3. Model estimates for Ab levels in immunosuppressed compared to immunocompetent

**A Analysis with all Ab data**

| Model variable | | Estimate | SE | p-value* |
|---|---|---|---|---|
| Intercept | | 3.095 | 0.265 | <0.0001 |
| Immunocompetent | TRUE | ref. | | |
| | FALSE | -0.807 | 0.103 | <0.0001 |
| Doses | 2 | ref. | | |
| | 3 | 0.287 | 0.118 | 0.016 |
| DaysPostLastDoseGroup | <2m | ref. | | |
| | 2-6m | -0.411 | 0.122 | 0.0010 |
| | >6m | -1.231 | 0.219 | <0.0001 |

**B Sensitivity analysis excluding censored Ab levels**

| Model variable | | Estimate | SE | p-value |
|---|---|---|---|---|
| Intercept | | 3.115 | 0.229 | <0.0001 |
| Immunocompetent | TRUE | ref. | | |
| | FALSE | -0.606 | 0.084 | <0.0001 |
| Doses | 2 | ref. | | |
| | 3 | 0.370 | 0.100 | 0.0003 |
| DaysPostLastDoseGroup | <2m | ref. | | |
| | 2-6m | -0.465 | 0.097 | <0.0001 |
| | >6m | -1.121 | 0.178 | <0.0001 |

Model: $\log_{10}$(Ab) ~ immunocompetent + Doses + DaysPostLastDoseGroup + (1 | Study)
Model fitting: weighting by study size, linear mixed model fit by REML (using 'lmer' from the lme4 package[32]).
* P-value for t-test using Satterthwaites's method ('lmerTest' package).

#### Table S4. Impact of treatments and conditions on Ab levels

| | | Estimate | SE | p-value* | ANOVA p-value† | LRT model comparison# |
|---|---|---|---|---|---|---|
| **1. *Doses + DaysPostLastDoseGroup*:** *AICc = 486.3* | | | | | | |
| Intercept | | 2.674 | 0.265 | <0.0001 | | - |
| Doses | 2 | ref. | | | 0.13 | |
| | 3 | 0.212 | 0.138 | 0.13 | | |
| DaysPostLastDoseGroup | <2m | ref. | | | <0.0001 | |
| | 2-6m | -0.438 | 0.143 | 0.0025 | | |
| | >6m | -1.269 | 0.255 | <0.0001 | | |
| **2. *Immunocompetent + Doses + DaysPostLastDoseGroup:*** *AICc = 435.6* | | | | | | |
| Intercept | | 3.096 | 0.258 | <0.0001 | | Comparison with model 1: p < 0.0001 |
| Immunocompetent | TRUE | ref. | | | <0.0001 | |
| | FALSE | -0.807 | 0.102 | <0.0001 | | |
| Doses | 2 | ref. | | | 0.015 | |
| | 3 | 0.287 | 0.117 | 0.015 | | |
| DaysPostLastDoseGroup | <2m | ref. | | | <0.0001 | |
| | 2-6m | -0.411 | 0.121 | 0.0008 | | |
| | >6m | -1.230 | 0.216 | <0.0001 | | |

| | | | | | | |
|---|---|---|---|---|---|---|
| **3. *Treatment + Doses + DaysPostLastDoseGroup*:** *AICc = 312.9* | | | | | | |
| Intercept | | 3.157 | 0.229 | <0.0001 | | Comparison with model 1: p < 0.0001 |
| Treatment | Untreated | ref. | | | <0.0001 | |
| | B-cell depletion | -1.737 | 0.098 | <0.0001 | | |
| | csDMARDs | -0.210 | 0.117 | 0.074 | | |
| | anti-IL6R | -0.519 | 0.171 | 0.0028 | | |
| | Integrin inhibitor | -0.071 | 0.187 | 0.70 | | |
| | JAKi | -0.581 | 0.162 | 0.0005 | | |
| | S1P inhibitors | -1.395 | 0.135 | <0.0001 | | |
| | anti-TNFα | -0.421 | 0.107 | 0.0001 | | |
| Doses | 2 | ref. | | | <0.0001 | |
| | 3 | 0.333 | 0.076 | <0.0001 | | |
| DaysPostLastDoseGroup | <2m | ref. | | | <0.0001 | |
| | 2-6m | -0.482 | 0.078 | <0.0001 | | |
| | >6m | -1.244 | 0.140 | <0.0001 | | |
| **4. *Condition + Doses + DaysPostLastDoseGroup*:** *AICc = 409.8* | | | | | | |
| Intercept | | 3.143 | 0.247 | <0.0001 | | Comparison with model 1: p < 0.0001<br><br>Comparison with model 2: p < 0.0001 |
| Condition | Healthy | ref. | | | <0.0001 | |
| | Haem | -0.689 | 0.255 | 0.0077 | | |
| | mixed IMIDs | -0.216 | 0.206 | 0.30 | | |
| | Neur | -1.536 | 0.157 | <0.0001 | | |
| | Rheu | -0.483 | 0.153 | 0.0019 | | |
| Doses | 2 | ref. | | | 0.0044 | |
| | 3 | 0.306 | 0.106 | 0.0044 | | |
| DaysPostLastDoseGroup | <2m | ref. | | | <0.0001 | |
| | 2-6m | -0.404 | 0.109 | 0.0003 | | |
| | >6m | -1.154 | 0.196 | <0.0001 | | |
| **5. *Treatment + Condition + Doses + DaysPostLastDoseGroup*:** *AICc = 315.7* | | | | | | |
| Intercept | | 3.166 | 0.229 | <0.0001 | | Comparison with model 1: p < 0.0001<br><br>Comparison with model 2: p < 0.0001<br><br>Comparison with model 3: p = 0.26<br><br>Comparison with model 4: p < 0.0001 |
| Treatment | Untreated | ref. | | | <0.0001 | |
| | B-cell depletion | -1.687 | 0.185 | <0.0001 | | |
| | csDMARDs | -0.343 | 0.213 | 0.11 | | |
| | anti-IL6R | -0.628 | 0.245 | 0.011 | | |
| | Integrin inhibitor | -0.042 | 0.239 | 0.86 | | |
| | JAKi | -0.661 | 0.235 | 0.0055 | | |
| | S1P inhibitors | -1.255 | 0.219 | <0.0001 | | |
| | anti-TNFα | -0.539 | 0.200 | 0.0080 | | |
| Condition | Healthy | ref. | | | 0.26 | |
| | Haem | -0.001 | 0.227 | 0.996 | | |
| | mixed IMIDs | 0.232 | 0.218 | 0.29 | | |
| | Neur | -0.229 | 0.209 | 0.28 | | |
| | Rheu | 0.163 | 0.215 | 0.45 | | |
| Doses | 2 | ref. | | | <0.0001 | |
| | 3 | 0.334 | 0.075 | <0.0001 | | |
| DaysPostLastDoseGroup | <2m | ref. | | | <0.0001 | |
| | 2-6m | -0.470 | 0.077 | <0.0001 | | |
| | >6m | -1.222 | 0.139 | <0.0001 | | |

* P-value for t-test using Satterthwaite's method ('lmerTest' package).
† Type III Analysis of Variance Table with Satterthwaite's method ('anova' function from the 'lmerTest' package).
### Likelihood ratio test comparison with another model (using the function 'anova' with test="LRT").
Model: $\log_{10}$(Ab) ~ *fixed effect(s)* + (1 | Study), where the fixed effects were Doses + DaysPostLastDoseGroup and treatment, condition, or both treatment and condition depending on the model.
Model fitting: weighting by study size, linear mixed model fit with REML (using lmer).

#### Table S5. Impact of treatments and conditions on Ab levels after removing censored Ab levels

| | | Estimate | SE | p-value* | ANOVA p-value† | LRT model comparison# |
|---|---|---|---|---|---|---|
| **1. *Doses + DaysPostLastDoseGroup*:** *AICc = 359.8* | | | | | | |
| Intercept | | 2.825 | 0.232 | <0.0001 | | - |
| Doses | 2 | ref. | | | 0.013 | |
| | 3 | 0.295 | 0.117 | 0.013 | | |
| DaysPostLastDoseGroup | <2m | ref. | | | <0.0001 | |
| | 2-6m | -0.470 | 0.114 | <0.0001 | | |
| | >6m | -1.129 | 0.210 | <0.0001 | | |
| **2. *Immunocompetent + Doses + DaysPostLastDoseGroup*:** *AICc = 321.1* | | | | | | |
| Intercept | | 3.115 | 0.229 | <0.0001 | | Comparison with model 1: $p < 0.0001$ |
| Immunocompetent | TRUE | ref. | | | <0.0001 | |
| | FALSE | -0.606 | 0.084 | <0.0001 | | |
| Doses | 2 | ref. | | | 0.0003 | |
| | 3 | 0.370 | 0.100 | 0.0003 | | |
| DaysPostLastDoseGroup | <2m | ref. | | | <0.0001 | |
| | 2-6m | -0.465 | 0.097 | <0.0001 | | |
| | >6m | -1.121 | 0.178 | <0.0001 | | |
| **3. *Treatment + Doses + DaysPostLastDoseGroup*:** *AICc = 250.1* | | | | | | |
| Intercept | | 3.154 | 0.216 | <0.0001 | | Comparison with model 1: $p < 0.0001$ |
| Treatment | Untreated | ref. | | | <0.0001 | |
| | B-cell depletion | -1.286 | 0.112 | <0.0001 | | |
| | csDMARDs | -0.226 | 0.102 | 0.029 | | |
| | anti-IL6R | -0.531 | 0.148 | 0.0005 | | |
| | Integrin inhibitor | 0.013 | 0.157 | 0.93 | | |
| | JAKi | -0.608 | 0.138 | <0.0001 | | |
| | S1P inhibitors | -1.372 | 0.126 | <0.0001 | | |
| | anti-TNFα | -0.433 | 0.094 | <0.0001 | | |
| Doses | 2 | ref. | | | <0.0001 | |
| | 3 | 0.418 | 0.070 | <0.0001 | | |
| DaysPostLastDoseGroup | <2m | ref. | | | <0.0001 | |
| | 2-6m | -0.486 | 0.067 | <0.0001 | | |
| | >6m | -1.139 | 0.126 | <0.0001 | | |

| | | | | | | |
|---|---|---|---|---|---|---|
| **4. *Condition* + *Doses* + *DaysPostLastDoseGroup***: *AICc = 309.7* | | | | | | |
| Intercept | | 3.160 | 0.232 | <0.0001 | | Comparison with model 1: $p < 0.0001$<br><br>Comparison with model 2: $p < 0.0001$ |
| Condition | Healthy | ref. | | | <0.0001 | |
| | Haem | -0.642 | 0.206 | 0.0022 | | |
| | mixed IMIDs | -0.220 | 0.164 | 0.18 | | |
| | Neur | -1.158 | 0.142 | <0.0001 | | |
| | Rheu | -0.383 | 0.128 | 0.0034 | | |
| Doses | 2 | ref. | | | <0.0001 | |
| | 3 | 0.384 | 0.093 | <0.0001 | | |
| DaysPostLastDoseGroup | <2m | ref. | | | <0.0001 | |
| | 2-6m | -0.483 | 0.089 | <0.0001 | | |
| | >6m | -1.089 | 0.165 | <0.0001 | | |
| **5. *Treatment* + *Condition* + *Doses* + *DaysPostLastDoseGroup***: *AICc = 263.5* | | | | | | |
| Intercept | | 3.160 | 0.218 | <0.0001 | | Comparison with model 1: $p < 0.0001$<br><br>Comparison with model 2: $p < 0.0001$<br><br>Comparison with model 3: $p = 0.44$<br><br>Comparison with model 4: $p < 0.0001$ |
| Treatment | Untreated | ref. | | | <0.0001 | |
| | B-cell depletion | -1.304 | 0.173 | <0.0001 | | |
| | csDMARDs | -0.438 | 0.199 | 0.029 | | |
| | anti-IL6R | -0.716 | 0.223 | 0.0017 | | |
| | Integrin inhibitor | -0.044 | 0.205 | 0.83 | | |
| | JAKi | -0.722 | 0.208 | 0.0007 | | |
| | S1P inhibitors | -1.348 | 0.203 | <0.0001 | | |
| | anti-TNF$\alpha$ | -0.626 | 0.185 | 0.0010 | | |
| Condition | Healthy | ref. | | | 0.50 | |
| | Haem | -0.043 | 0.195 | 0.82 | | |
| | mixed IMIDs | 0.258 | 0.191 | 0.18 | | |
| | Neur | -0.056 | 0.192 | 0.77 | | |
| | Rheu | 0.226 | 0.201 | 0.26 | | |
| Doses | 2 | ref. | | | <0.0001 | |
| | 3 | 0.416 | 0.071 | <0.0001 | | |
| DaysPostLastDoseGroup | <2m | ref. | | | <0.0001 | |
| | 2-6m | -0.488 | 0.067 | <0.0001 | | |
| | >6m | -1.136 | 0.126 | <0.0001 | | |

* P-value for t-test using Satterthwaite's method ('lmerTest' package).

† Type III Analysis of Variance Table with Satterthwaite's method ('anova' function from the 'lmerTest' package).

### Likelihood ratio test comparison with another model (using the function 'anova' with test="LRT").

Model: $\log_{10}$(Ab) ~ *fixed effect(s)* + (1 | Study), where the fixed effects were Doses + DaysPostLastDoseGroup and treatment, condition, or both treatment and condition depending on the model.

Model fitting: weighting by study size, linear mixed model fit with REML (using lmer).

Data: Binding Ab level data excluding censored Ab levels (i.e. Ab levels reported at the lower or upper limit of quantification; Table S1).

#### Table S6. Ab data – model selection by lowest AICc

##### A Analysis with all Ab data

| | |
|---|---|
| **Possible fixed effects** | Vaccine, Doses, DaysPostLastDoseGroup, Response, Treatment, Condition, immunocompetent |
| **Best model fixed effects** | 1 + Treatment + Doses + DaysPostLastDoseGroup |
| **AICc** | 315.3 |
| **Number of parameters** | 11 |

##### B Sensitivity analysis excluding censored Ab levels

| | |
|---|---|
| **Possible fixed effects** | Vaccine, Doses, DaysPostLastDoseGroup, Response, Treatment, Condition, immunocompetent |
| **Best model fixed effects** | 1 + Treatment + Doses + DaysPostLastDoseGroup |
| **AICc** | 221.9 |
| **Number of parameters** | 11 |

Models: $\log_{10}$(Ab) ~ *fixed effect(s)* + (1 | Study).
Model fitting: weighting by study size, linear mixed model fit (using lmer with REML = FALSE). Note that statistical models were fit using REML = FALSE for model comparison but REML = TRUE for model outputs (Table S4 and Table S5).

#### Table S7. Ratio of Ab levels between treatments and conditions

##### A Treatment comparisons

| | Untreat-ed | B-cell depl. | S1P inh. | JAKi | anti-IL6R | anti-TNFα | csDMARDs | Integrin inh. |
|---|---|---|---|---|---|---|---|---|
| **Untreated** | | 54.536 | 24.827 | 3.812 | 3.302 | 2.637 | 1.624 | 1.178 |
| **B-cell depl.** | 0.018 | | 0.455 | 0.070 | 0.061 | 0.048 | 0.030 | 0.022 |
| **S1P inh.** | 0.040 | 2.197 | | 0.154 | 0.133 | 0.106 | 0.065 | 0.047 |
| **JAKi** | 0.262 | 14.308 | 6.514 | | 0.866 | 0.692 | 0.426 | 0.309 |
| **anti-IL6R** | 0.303 | 16.518 | 7.520 | 1.155 | | 0.799 | 0.492 | 0.357 |
| **anti-TNFα** | 0.379 | 20.680 | 9.415 | 1.445 | 1.252 | | 0.616 | 0.447 |
| **csDMARDs** | 0.616 | 33.583 | 15.288 | 2.347 | 2.033 | 1.624 | | 0.725 |
| **Integrin inh.** | 0.849 | 46.305 | 21.080 | 3.236 | 2.803 | 2.239 | 1.379 | |

##### B Condition comparisons

| | Healthy | Neur | Haem | Rheu | mixed IMIDs |
|---|---|---|---|---|---|
| **Healthy** | | 34.366 | 4.896 | 3.038 | 1.646 |
| **Neur** | 0.029 | | 0.142 | 0.088 | 0.048 |
| **Haem** | 0.204 | 7.019 | | 0.620 | 0.336 |
| **Rheu** | 0.329 | 11.312 | 1.612 | | 0.542 |
| **mixed IMIDs** | 0.608 | 20.881 | 2.975 | 1.846 | |

Treatments or conditions are compared using contrast analysis (with the function 'contest'). The ratio of Ab levels is for the treatment or condition in the row compared to the column. Grey shading indicates significant differences ($p<0.05$).

#### Table S8. Ratio of Ab levels between treatments and conditions after removing censored Ab levels

**A Treatment comparisons**

| | Untreat-ed | B-cell depl. | S1P inh. | JAKi | anti-IL6R | anti-TNFα | csDMARDs | Integrin inh. |
|---|---|---|---|---|---|---|---|---|
| **Untreated** | | 19.332 | 23.544 | 4.057 | 3.396 | 2.713 | 1.684 | 0.969 |
| **B-cell depl.** | 0.052 | | 1.218 | 0.210 | 0.176 | 0.140 | 0.087 | 0.050 |
| **S1P inh.** | 0.042 | 0.821 | | 0.172 | 0.144 | 0.115 | 0.072 | 0.041 |
| **JAKi** | 0.246 | 4.765 | 5.803 | | 0.837 | 0.669 | 0.415 | 0.239 |
| **anti-IL6R** | 0.294 | 5.693 | 6.932 | 1.195 | | 0.799 | 0.496 | 0.285 |
| **anti-TNFα** | 0.369 | 7.127 | 8.679 | 1.496 | 1.252 | | 0.621 | 0.357 |
| **csDMARDs** | 0.594 | 11.478 | 13.978 | 2.409 | 2.016 | 1.610 | | 0.576 |
| **Integrin inh.** | 1.031 | 19.941 | 24.285 | 4.185 | 3.503 | 2.798 | 1.737 | |

**B Condition comparisons**

| | Healthy | Neur | Haem | Rheu | mixed IMIDs |
|---|---|---|---|---|---|
| **Healthy** | | 14.389 | 4.389 | 2.416 | 1.661 |
| **Neur** | 0.069 | | 0.305 | 0.168 | 0.115 |
| **Haem** | 0.228 | 3.279 | | 0.550 | 0.378 |
| **Rheu** | 0.414 | 5.956 | 1.817 | | 0.688 |
| **mixed IMIDs** | 0.602 | 8.663 | 2.642 | 1.454 | |

Treatments or conditions are compared using contrast analysis (with the function 'contest'). The ratio of Ab levels is for the treatment or condition in the row compared to the column. Grey shading indicates significant differences ($p<0.05$).

Models: $\log_{10}$(Ab) ~ Treatment + Doses + DaysPostLastDoseGroup + (1 | Study) or $\log_{10}$(Ab) ~ Condition + Doses + DaysPostLastDoseGroup + (1 | Study), for treatments and conditions, respectively (see Table S5).

Data: Binding Ab level data excluding censored Ab levels (i.e. Ab levels reported at the lower or upper limit of quantification; Table S1).

#### Table S9. Vaccine effectiveness (VE) studies and linking method to Ab data

| Study | Vaccines | Doses (time since last dose) | Variant | Conditions | Treatments | Linked by |
|---|---|---|---|---|---|---|
| Britton[33] | mRNA | 3 (≥7 days) | Omicron (BA.1 to BA.5) | Haem (hematologic malignancies) | Mixed/unknown (no information) | Condition |
| | | 2 (≥14 days), 3 (≥7 days) | | mixed IMIDs (rheumatologic or inflammatory disorder) | | |
| Calabrese[34] | mRNA | 1-3 | pre-Omicron | mixed IMIDs (immune-mediated inflammatory diseases) | B-cell depletion (+ DMARDs for 17.6% in the breakthrough cohort) | Treatment |

| | | | | | | |
|---|---|---|---|---|---|---|
| | (in the breakthrough cohort: 91.9% mRNA, 8.1% J&J) | | | | | |
| Desai[35] | mRNA (Pfizer 80.6%, Moderna 19.4%) | 1, 2, 3, 1-3 | pre-Omicron to Omicron (to BA.5) | mixed IMIDs (IBD) | Mixed/unknown (mesalamine, TNFi, immunomodulator, vedolizumab, ustekinumab, tofacitinib) | Condition |
| Embi (1)[36] | Moderna, Pfizer | 2 | pre-Omicron (pre-Delta and Delta) | Haem (hematologic malignancies) | Mixed/unknown (no information) | Condition |
| | | | | mixed IMIDs (rheumatologic or inflammatory disorder) | | |
| Embi (2)[37] | mRNA | 2, 3 | pre-Omicron (Delta) | Haem (hematologic malignancies only) | Mixed/unknown (no information) | Condition |
| | | 2, 3 | | mixed IMIDs (rheumatologic or inflammatory disorder only) | | |
| Gazitt[38] | mRNA | 1, 2, 3 | pre-Omicron | mixed IMIDs (psoriatic disease) | Mixed/unknown (no information) | Condition |
| Khan (1)[39] | mRNA | 1, 2 | pre-Omicron (Ancestral, Alpha, Beta, Gamma) | mixed IMIDs (IBD) | csDMARDs, integrin inhibitor, anti-TNFα, and JAKi, others (~1%) | Treatments |
| Khan (2)[40] | mRNA (98.5% mRNA, 1.5% J&J and AZ) | 2, 3 | Omicron (BA.1) | mixed IMIDs (IBD) | csDMARDs, anti-TNFα, JAKi, integrin inhibitor, others (<3%) | Treatments |
| Smith[41] | mRNA (93.5% mRNA in total cohort) | 2-3 (99.4% 2 or 3 doses in total cohort, 0.5% 4 doses, 0.1% 5 doses) | pre-Omicron to Omicron (Alpha to Omicron BA.1) | Neur (multiple sclerosis) | B-cell depletion (rituximab) | Treatment |
| Spierer[42] | Pfizer | 2 | pre-Omicron (Alpha) | Neur (multiple sclerosis) | Mixed/unknown (cladribine, ocrelizumab, natalizumab, S1P receptor modulator, interferon beta, others) | Condition |

| | | | | | | |
|---|---|---|---|---|---|---|
| Widdifield[43] | mRNA | 1, 2, 3 | pre-Omicron (Alpha, Delta) | Rheu (rheumatoid arthritis) | Mixed/unknown (no information) | Condition |
| | | 1, 2, 3 | | Rheu (ankylosing spondylitis) | | |
| | | 1, 2, 3 | | mixed IMIDs (psoriasis) | | |
| | | 1, 2, 3 | | mixed IMIDs (IBD) | | |
| Wu[44] | mRNA (99% mRNA, 0.1% unknown) | 1, 2 | pre-Omicron (Ancestral to Beta) | Haem (hematologic cancer) | Mixed (chemotherapy-containing, targeted treatment, endocrine treatment) | Condition |

#### Table S10. Univariate meta-analysis of VE by $log_{10}$-Ab levels

**A Analysis with all Ab data**

| | Estimate | SE | p-value† | $I^2$ (95% CI) |
|---|---|---|---|---|
| ***Outcome infection:*** | | | | |
| Intercept<br>$log_{10}$(Ab) | -1.019<br>-0.775 | 0.159<br>0.047 | <0.0001<br><0.0001 | 94.10<br>(83.80-98.48) |
| ***Outcome hospitalization:*** | | | | |
| Intercept<br>$log_{10}$(Ab) | -1.467<br>-0.780 | 0.209<br>0.108 | <0.0001<br><0.0001 | 86.89<br>(69.97-95.96) |
| ***Outcome death:*** | | | | |
| Intercept<br>$log_{10}$(Ab) | -0.919<br>-1.158 | 0.291<br>0.433 | 0.0016<br>0.0075 | 15.44<br>(0-92.50) |

**B Sensitivity analysis excluding censored Ab levels**

| | Estimate | SE | p-value | $I^2$ (95% CI) |
|---|---|---|---|---|
| ***Outcome infection:*** | | | | |
| Intercept<br>$log_{10}$(Ab) | -0.951<br>-0.802 | 0.142<br>0.048 | <0.0001<br><0.0001 | 92.46<br>(80.47-98.02) |
| ***Outcome hospitalization:*** | | | | |
| Intercept<br>$log_{10}$(Ab) | -1.447<br>-0.830 | 0.188<br>0.111 | <0.0001<br><0.0001 | 83.93<br>(64.69-94.95) |
| ***Outcome death:*** | | | | |
| Intercept<br>$log_{10}$(Ab) | -0.915<br>-1.156 | 0.292<br>0.436 | 0.0018<br>0.0080 | 17.01<br>(0-92.71) |

† P-value of for the test of individual coefficients in the model (based on a standard normal distribution, from ‘rma.mv’ function).
Model: log(RR) ~ $log_{10}$(Ab) + (1 | Study), where RR = 1-VE/100.
Model fitting: using rma.mv (from the metafor package[45]) with inverse variance weighting and method ‘REML’.
Data: all VE data that could be linked with an Ab level (see Table S9), stratified by outcome (infection, hospitalization, death).

#### Table S11. VE data with Ab levels – model selection by lowest AICc

**A Analysis with all Ab data**

| Outcome | Infection | Hospitalization |
| --- | --- | --- |
| **Possible moderators** | StudyType, Vaccine, Treatment, Condition, Doses, VariantGroup, Omicron, effect measure, $\log_{10}$(Ab) | |
| **Best model** | yi ~ 1 + Doses + VariantGroup + StudyType + $\log_{10}$(Ab) | yi ~ 1 + Doses + Treatment + Omicron + effect measure |
| **AICc** | 182.0 | 56.4 |
| **$I^2$ (95% CI)** | 0% (0 – 16.73) | 82.13% (55.41 – 96.73) |
| **Test for Residual Heterogeneity** | QE(df = 17) = 193.30 $p < 0.0001$ | QE(df = 23) = 85.36 $p < 0.0001$ |
| **Test of Moderators** | QM(df = 8) = 587.50 $p < 0.0001$ | QM(df = 7) = 88.69 $p < 0.0001$ |

**B Sensitivity analysis excluding censored Ab levels**

| Outcome | Infection | Hospitalization |
| --- | --- | --- |
| **Possible moderators** | StudyType, Vaccine, Treatment, Condition, Doses, VariantGroup, Omicron, effect measure, $\log_{10}$(Ab) | |
| **Best model** | yi ~ 1 + Doses + Omicron + StudyType + $\log_{10}$(Ab) | yi ~ 1 + Doses + effect measure + $\log_{10}$(Ab) |
| **AICc** | 179.2 | 55.6 |
| **$I^2$ (95% CI)** | 52.2% (0 – 99.1) | 83.3% (57.80 – 96.41) |
| **Test for Residual Heterogeneity** | QE(df = 17) = 190.83 $p < 0.0001$ | QE(df = 23) = 82.37 $p < 0.0001$ |
| **Test of Moderators** | QM(df = 8) = 379.22 $p < 0.0001$ | QM(df = 7) = 90.02 $p < 0.0001$ |

Model: log(RR) ~ *moderator(s)* + (1 | Study), where RR = 1-VE/100 and all possible moderators are listed in the Methods section (Model comparison, pp. 7-8).
Model fitting: using rma.mv (from the metafor package[45]) with inverse variance weighting and method 'ML' (due to model comparisons). Models are compared using AICc.
Data: all VE data that could be linked with an Ab level, stratified by outcome (infection and hospitalization; death outcome was excluded due to limited available data, Fig. S5 and Fig. S8C).

#### Table S12. VE data with Ab levels – lowest AICc models for outcomes infection and hospitalization

| | | Estimate | SE | p-value[†] |
|---|---|---|---|---|
| ***Outcome infection:*** *AICc = 224.8* | | | | |
| Intercept | | -0.987 | 0.031 | <0.0001 |
| $log_{10}$(Ab) | | -0.363 | 0.153 | 0.018 |
| Doses | 1 | 0.722 | 0.107 | <0.0001 |
| | 2 | ref. | | |
| | 3 | -0.967 | 0.119 | <0.0001 |
| | 1-3 | 1.204 | 0.273 | <0.0001 |
| VariantGroup | pre-Omicron | ref. | | |
| | pre-Omicron to Omicron | 0.654 | 0.154 | <0.0001 |
| | Omicron BA.1 | 1.055 | 0.185 | <0.0001 |
| StudyType | Nested matched case-control | ref. | | |
| | Retrospective cohort | 0.415 | 0.152 | 0.0062 |
| | Test-negative case-control | -0.449 | 0.045 | <0.0001 |
| ***Outcome hospitalization:*** *AICc = 57.2* | | | | |
| Intercept | | 0.661 | 0.653 | 0.31 |
| Doses | 1 | 1.101 | 0.150 | <0.0001 |
| | 2 | ref. | | |
| | 3 | -0.493 | 0.116 | <0.0001 |
| | 1-3 | 1.435 | 0.590 | 0.015 |
| | 2-3 | 0.432 | 0.645 | 0.50 |
| Treatment | Mixed/unknown | ref. | | |
| | csDMARDs, Integrin inhibitors, JAKi, anti-anti-TNFα | -1.892 | 0.793 | 0.017 |
| | B-cell depletion | * | | |
| Omicron | pre-Omicron | ref. | | |
| | pre-Omicron to Omicron | * | | |
| | Omicron | 1.197 | 0.482 | 0.013 |
| Effect measure | aHR | ref. | | |
| | aOR | -2.496 | 0.608 | <0.0001 |
| | OR | * | | |

[†] P-value of for the test of individual coefficients in the model (based on a standard normal distribution, from 'rma.mv' function).
* Redundant predictor dropped from the model.
Model: log(RR) ~ *moderator(s)* + (1 | Study), where RR = 1-VE/100 and are the moderators for the best model for each outcome (by AICc, see Table S11).
Model fitting: using rma.mv (from the metafor package[45]) with inverse variance weighting and method 'REML'.
Data: all VE data that could be linked with an Ab level, stratified by outcome (infection and hospitalization; death outcome was excluded due to limited available data, Fig. S5).

#### Table S13. VE data with Ab levels – lowest AICc models with Ab levels excluding censored data

| | | Estimate | SE | p-value† |
|---|---|---|---|---|
| ***Outcome infection:*** *AICc = 222.0* | | | | |
| Intercept | | -0.509 | 0.170 | 0.0028 |
| $\log_{10}$(Ab) | | 0.623 | 0.212 | 0.0033 |
| Doses | 1 | 0.893 | 0.144 | <0.0001 |
| | 2 | ref. | | |
| | 3 | -1.138 | 0.153 | <0.0001 |
| | 1-3 | 1.246 | 0.299 | <0.0001 |
| Omicron | pre-Omicron | ref. | | |
| | pre-Omicron to Omicron | 0.700 | 0.211 | 0.0009 |
| | Omicron | 1.258 | 0.258 | <0.0001 |
| StudyType | Nested matched case-control | -0.491 | 0.214 | 0.022 |
| | Retrospective cohort | ref. | | |
| | Test-negative case-control | -0.940 | 0.210 | <0.0001 |
| ***Outcome hospitalization:*** *AICc = 56.9* | | | | |
| Intercept | | -0.965 | 0.454 | 0.034 |
| $\log_{10}$(Ab) | | -0.625 | 0.229 | 0.0064 |
| Doses | 1 | 0.679 | 0.206 | 0.0010 |
| | 2 | ref. | | |
| | 3 | -0.028 | 0.198 | 0.89 |
| | 1-3 | 1.131 | 0.605 | 0.061 |
| | 2-3 | -0.290 | 0.680 | 0.67 |
| Effect measure | aHR | ref. | | |
| | aOR | -0.774 | 0.501 | 0.12 |
| | OR | 1.624 | 0.784 | 0.038 |

† P-value of for the test of individual coefficients in the model (based on a standard normal distribution, from 'rma.mv' function).

Model: log(RR) ~ *moderator(s)* + (1 | Study), where RR = 1-VE/100 and are the moderators for the best model for each outcome (by AICc, see Table S11).

Model fitting: using rma.mv (from the metafor package[45]) with inverse variance weighting and method 'REML'.

Data: all VE data that could be linked with an Ab level, stratified by outcome (infection and hospitalization; death outcome was excluded due to limited available data, Fig. S8C). Binding Ab level data excludes censored Ab levels (i.e. Ab levels reported at the lower or upper limit of quantification; Table S1) and was used to adjust for immunosuppressive treatment or underlying condition.

### Abbreviations

| | |
|---|---|
| AAV | antineutrophil cytoplasmic antibody-associated vasculitis |
| Ab | antibody |
| aHR | adjusted hazard ratio |
| AICc | Akaike information criterion with small sample size correction |
| AIHA | autoimmune hemolytic anemia |
| ANOVA | analysis of variance |
| anti-IL6R | anti-IL6 receptor |
| anti-TNF$\alpha$ | anti-tumor necrosis factor alpha |
| aOR | adjusted odds ratio |
| AUC | area under the curve |
| AU/mL | arbitrary units per milliliter |
| AxSpA | axial spondyloarthritis |
| AZ (AstraZeneca) | Oxford-AstraZeneca COVID-19 vaccine (ChAdOx1 nCoV-19) |
| BAU/mL | binding antibody units per milliliter |
| BTKi | Bruton-kinase inhibitor |
| CLIA | chemiluminescent immunoassay |
| CMIA | chemiluminescent microparticle immunoassay |
| csDMARDs | conventional synthetic disease modifying anti-rheumatic drugs |
| CTD | connective tissue disease |
| ECLIA | electrochemiluminescence sandwich immunoassay |
| ELISA | enzyme-linked immunosorbent assay |
| ELISpot | enzyme-linked immunosorbent spot |
| FEIA | fluorescence enzyme immunoassay |
| Haem | hematological malignancies |
| HR | hazard ratio |
| $I^2$ | statistic describing the percent of variation that is due to study heterogeneity |
| IBD | inflammatory bowel disease |
| IgG | immunoglobulin G |
| inh. | inhibitor |
| IMIDs | immune-mediated inflammatory disorders |
| IQR | interquartile range |
| IT | immune thrombocytopenia |
| JAKi | JAK inhibitor |
| J&J | Johnson & Johnson COVID-19 vaccine (Ad26.COV2.S) |
| LRT | likelihood ratio test |
| LVV | large vessel vasculitis |
| ML | maximum likelihood |
| MPD | myeloproliferative disease |
| MS | multiple sclerosis |
| N | number of study participants contributing to the data |
| Neur | neurological disorders |
| NHL | non-Hodgkin’s lymphoma |
| OR | odds ratio |

| | |
|---|---|
| PsA | psoriatic arthritis |
| R | range |
| RA | rheumatoid arthritis |
| RBD | receptor binding domain |
| ref. | reference |
| REML | restricted maximum likelihood |
| Rheu | rheumatic diseases |
| RR | relative risk |
| RU/mL | relative units per milliliter |
| S1 | S1 subunit of the SARS-CoV-2 Spike Protein |
| S1P inhibitor | sphingosine-1-phosphate inhibitor |
| SE | standard error |
| SEM | standard error of the mean |
| SLE | systematic lupus erythematosus |
| TSP | trimeric spike protein |
| U/mL | standardized units per milliliter |
| VE | vaccine effectiveness |
